# Cell-type-specific Transcriptomic-wide Association Studies Detected 91 Independent Risk Genes for Alzheimer’s Disease Dementia

**DOI:** 10.1101/2025.05.27.25328452

**Authors:** Qiang Liu, Randy L. Parrish, Shizhen Tang, Shinya Tasaki, David A. Bennett, Nicholas T. Seyfried, Philip L. De Jager, Vilas Menon, Aron S. Buchman, Jingjing Yang

## Abstract

Existing TWASs of AD dementia typically use a single statistical method to identify cell-type-specific risk genes. Here we sought to improve on existing approaches and utilized an omnibus xWAS pipeline to integrate snRNA-seq dataset (n=415) of dorsolateral prefrontal cortex (DLPFC) and the latest GWAS data of AD dementia to detect cell-type-specific risk genes. We identified 223 cell-type-specific TWAS risk genes across six major brain cell types, including 91 independent associations of which 11 are novel. Integrating proteomics data (n=716) of DLPFC and GWAS data, we identified 21 significant PWAS risk genes including 13 independent associations, overlapping with 32% independent cell-type-specific TWAS associations. Protein-protein interaction network analyses showed that our TWAS findings are functionally linked to established AD risk genes such as *APOE, BIN1*, and *MAPT*. These results underscore the value of leveraging large-scale snRNA-seq and proteomics data to uncover novel cell-type-specific mechanisms underlying AD dementia.

---

Alzheimer’s disease (AD) is the most common cause of dementia without effective cure, affecting millions of individuals worldwide^1^. Despite extensive research, the molecular mechanisms underlying AD remain largely unknown, highlighting the potential utility of integrative analyses of genetic, transcriptomic, and proteomic data to fill this critical knowledge gap. Recently, transcriptome-wide association studies^2^ (TWASs) and proteome-wide association studies^3^ (PWASs) have emerged as powerful tools for integrating transcriptomic and proteomic data with summary-level data by genome-wide association studies^4^ (GWASs). The standard two-stage TWAS/PWAS (i.e., xWAS) approach^5–8^ (see Methods) first estimates the cis-genetic effect sizes (referred to cis-xQTL weights) on each quantitative gene expression or protein abundance trait, and then takes these effect sizes as weights to conduct gene-based association test for each gene (or the corresponding protein coding gene). Genes with significant xWAS p-values have their genetic effects on the phenotype of interest in the GWAS data potentially mediated through their genetically regulated gene expressions or protein abundances.

Although recent TWASs^9–11^ of AD detected >100 risk genes including both known GWAS risk genes and novel findings, these analyses primarily studied bulk RNA sequencing (RNA-seq) data of AD-related tissues such as dorsolateral prefrontal cortex (DLPFC) and often only used one statistical method. Recent studies utilizing single-nucleus RNA sequencing (snRNA-seq, including pre-mRNA and nuclear-localized RNA) have revealed critical insights into cell-type-specific mechanisms underlying AD^12,13^. For example, genetic variants associated with AD were shown with cell-type-specific patterns of enhancer enrichment, particularly in microglia cells^14^, and expression quantitative trait loci (eQTL) in microglia cells are shown most enriched with GWAS risk loci of AD^15,16^. Also, recent snRNA-seq studies identified novel biomarkers, uncovered disease-associated pathways, and elucidate the important roles of non-neuronal cells such as microglia and astrocytes in AD pathologies^17^. These findings collectively highlight the importance of studies exploring cell-type-specific genomic mechanisms underlying AD.

Here, we applied our recently developed omnibus xWAS-O pipeline^18^ to integrate the latest large-scale snRNA-seq data of DLPFC (n=415) from the Religious Orders Study and Rush Memory and Aging Project (ROSMAP) Study^19^ and the latest GWAS summary data of AD (Bellenguez et. al 2022. N=∼789K)^4^. We studied six major brain cell types: astrocytes, microglia, excitatory neurons, inhibitory neurons, oligodendrocytes, and oligodendrocyte precursor cells.

In contrast to recent cell-type-specific TWAS analyses^19^ that used only one statistical method^8^ to model the genetic effect sizes on gene expressions, the xWAS-O pipeline^18^ aggregates TWAS findings based on multiple complementary statistical regression models (see Methods). In prior work, we have shown that this approach shows improved power and calibrated false positive rates^18,20^.

To calibrate for TWAS false positive rates, we further applied the recently developed Genomic Integration Fine-mapping Tool^21^ (GIFT) to fine map independent risk genes. Among 223 significant cell-type-specific TWAS risk genes, we fine-mapped 91 independent risk genes of which 11 were novel. Additionally, our PWAS-O analysis detected 21 significant PWAS risk genes, including 13 fine-mapped independent significant genes. We validated 29 of the 91 (32%) independent TWAS risk genes in the analogous PWAS-O findings using proteomics data from DLPFC (n=716). Further, protein-protein interaction (PPI) network analyses demonstrated that many of these cell-type-specific TWAS risk genes were interconnected with known AD risk genes such as *APOE*^22^, *BIN1*^23^, and *MAPT*^24^.

## Results

### Methods Overview

#### xWAS-O pipeline

As shown in **Fig. 1**, the xWAS-O pipeline^18^ is designed to integrate transcriptomic/proteomic data and GWAS summary data using multiple complementary statistical tools. Specifically, cix-xQTL weights are estimated in Stage I using the nonparametric Bayesian regression Dirichlet Process Regression (DPR) model implemented in TIGAR^5,6^, penalized regression model with Elastic-Net (EN) penalty implemented in PrediXcan^7^, as well as the best mode with the highest cross-validation (CV) *R*^2^ out of penalized regression models with EN and LASSO penalties, regular linear regression model with best unbiased linear predictor (BLUP), and single variant model with Top xQTL (Top 1) as implemented in FUSION^8^ (Methods).

**Figure 1.**
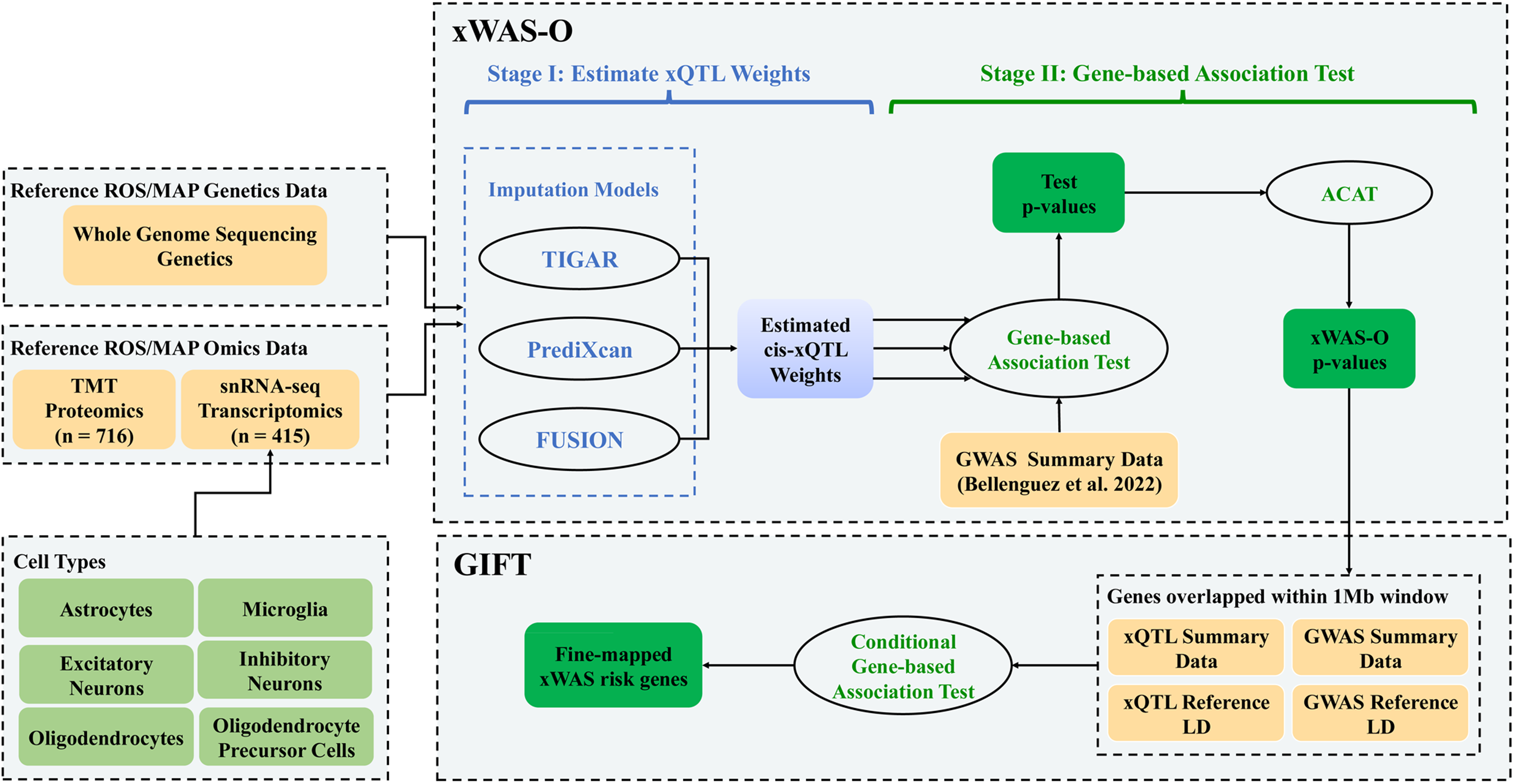
Overview of xWAS-O pipeline. Design of this study including inputs and outputs used in the xWAS-O pipeline.

For each gene, TWAS/PWAS p-values (Stage II) are obtained based on each set of the xQTL weights and GWAS summary data, which are then combined by the aggregated Cauchy association test (ACAT)^25^ to derive an omnibus p-value, referred to as xWAS-O p-values (Methods). In a prior PWAS-O study, we showed that this approach improved power and calibrated type I error under the null hypothesis^18^. Significant TWAS/PWAS risk genes are then reported based on xWAS-O p-values.

Since conventional TWAS/PWAS findings report inflated false positive rates^9–11^, the current study employed the recently proposed tool GIFT^21^ to further fine-map independent significant risk genes detected by xWAS-O (Methods). For each genetic region containing multiple nearby significant risk genes, we used GIFT to perform the analogous gene-level conditional association testing within a ±1Mb window around the top significant gene. Genes with independent significant associations are then reported.

#### ROS/MAP omics data

Here, we utilized large-scale, multi-omics data generated from the Religious Orders Study and Memory and Aging Project (ROS/MAP)^26^, which include single-nucleus RNA sequencing (snRNA-seq), proteomics, and genotype data from postmortem dorsolateral prefrontal cortex (DLPFC) tissue. The snRNA-seq data comprised over 1.5 million nuclei across six major brain cell types from 436 individuals, while the proteomic data included abundance profiles proteins from 716 individuals. Genotype data were profiled by whole genome sequencing (WGS)^27^. Expression and protein traits were normalized, log-transformed, and corrected for relevant biological and technical covariates prior to analysis (Methods). Only samples with both genotype and gene expression (or protein abundance) traits were used for estimating the corresponding cis-xQTL weights. As a result, the numbers of analyzed genes with pseudo-bulk gene expression across six brain cell types in 415 samples are presented in **Table 1**. Proteome abundance data of 8,929 proteins of 716 individuals were used for estimating cis-pQTL weights. These ROS/MAP reference snRNA-seq and proteomic datasets are one of the largest available omics datasets profiled from DLPFC, an AD-relevant tissue.

**Table 1:** The total number of test genes for studying AD dementia by three individual tools. Models of TIGAR/DPR, PrediXcan/EN, and FUSION/BestModel (first column) were applied to the ROS/MAP reference omics data (second column). Genes (or protein coding genes) with CV R2 > 0.5% for the corresponding gene expression (or protein abundance) imputation models were selected as test genes in PWAS/TWAS (third column). Median CV R2 of all test genes are presented in the fourth column.

| Model | Omics Data | Number of Test Genes | Median CV R<sup>2</sup> |
| --- | --- | --- | --- |
| TIGAR/DPR | Astrocytes | 13,482 | 0.013 |
|  | Excitatory neurons | 13,649 | 0.014 |
|  | Inhibitory neurons | 13,389 | 0.013 |
|  | Microglia | 13,458 | 0.013 |
|  | Oligodendrocytes | 13,462 | 0.013 |
|  | Oligodendrocyte precursor cells | 13,282 | 0.012 |
|  | Proteomic | 4,069 | 0.013 |
| PrediXcan/EN | Astrocytes | 4,536 | 0.012 |
|  | Excitatory neurons | 4,929 | 0.016 |
|  | Inhibitory neurons | 4,329 | 0.011 |
|  | Microglia | 4,126 | 0.011 |
|  | Oligodendrocytes | 4,155 | 0.012 |
|  | Oligodendrocyte precursor cells | 3,898 | 0.011 |
|  | Proteomic | 2,003 | 0.041 |
| FUSION/BestModel | Astrocytes | 5,478 | 0.0102 |
|  | Excitatory neurons | 5,478 | 0.0102 |
|  | Inhibitory neurons | 5,176 | 0.0099 |
|  | Microglia | 5,107 | 0.0095 |
|  | Oligodendrocytes | 5,138 | 0.0099 |
|  | Oligodendrocyte precursor cells | 4,710 | 0.0100 |
|  | Proteomic | 4,710 | 0.0192 |

#### Apply xWAS-O pipeline to ROS/MAP data

First, we applied the xWAS-O pipeline^18^ (**Fig. 1**) to ROS/MAP omics data to estimate cell-type-specific cis-eQTL and cis-pQTL weights by TIGAR/DPR, PrediXcan/EN, FUSION/BestModel. As suggested by TIGAR V2 paper^6^, only genes with CV *R*^2^ > 0.5% explained by estimated cis-xQTL weights underwent further testing for their association with AD. Second, xWAS p-values were obtained using each set of cis-xQTL weights and the latest GWAS summary data of AD dementia (n=∼789K)^4^, which were then used to obtain omnibus xWAS p-values by ACAT^25^. For each cell type, genes with omnibus TWAS p-values < 2.5E-06 are defined as significant TWAS-O findings. For proteome-wide analyses, we first adjusted the PWAS-O p-values according to the genomic control factor^28^ to ensure that the median observed PWAS-O p-value was adjusted to the expected value of 0.5 under the null hypothesis. From these adjusted nominal PWAS-O p-values, we then derived q-values to control for false discovery rates (FDRs) by Benjamini-Hochberg procedure^29^. Genes with PWAS-O q-values<0.05 are defined as significant PWAS-O findings, controlling the family-wise type I error rate at 0.05.

Third, for genomic regions containing multiple significant xWAS risk genes with overlapped test xQTL, we applied GIFT^21^ to fine map independent risk genes within ±1Mb window around the top significant gene. Independent significant risk genes include the ones either detected by GIFT with GIFT p-value<0.05, or having no overlapped test cis-xQTL with other significant risk genes.

Further, we compared cell-type-specific TWAS-O findings to the bulk TWAS-O and PWAS-O results (**Tables S1-S2**) that were respectively obtained by using the large-scale bulk RNA-seq data (n=931)^20^ and proteomics data (described above) of DLPFC in the same ROS/MAP cohort and the same GWAS summary statistics of AD dementia^4^. For each cell type, its cell-type-specific TWAS-O risk genes are considered as overlapping with bulk TWAS-O findings, when their test genetic regions overlapped. Independent cell-type-specific TWAS risk genes are considered as validated in PWAS-O findings, when their test genetic regions overlapped with any of the protein-coding genes with PWAS-O q-values<0.05.

#### PPI network analyses by STRING

We further provide the lists of significant cell-type-specific TWAS-O risk genes and PWAS-O risk genes to the STRING^30–32^ webtool, which generates protein-protein interaction (PPI) networks corresponding to the provided genes

#### Enrichment analyses by pathDIP

We also provide the lists of significant cell-type-specific TWAS-O risk genes and PWAS-O risk genes to the pathDIP^33^ webtool, which provides gene set enrichment analysis with databases of Panther^34^, PathBank^35^ and Reactome^36^.

### Cell-type-specific TWAS-O Results

As shown in **Table 1**, for each cell type, ∼13K valid gene imputation models with CV *R*^2^ > 0.5% were obtained by TIGAR/DPR with median CV *R*^2^= ∼0.013; ∼4K valid gene imputation models were obtained by PrediXcan/EN with median CV *R*^2^=∼0.012; ∼5K valid gene imputation models were obtained by FUSION/BestModel with median CV *R*^2^=∼0.01. Scatter plots and histograms of CV *R*^2^ for the three tools are presented in **Figs. S1-S3**. For each cell type, TWAS p-values were first obtained by using cis-eQTL weights estimated for transcriptome-wide genes with valid expression imputation models and the GWAS summary data of AD dementia^4^. Then TWAS-O p-values were obtained by ACAT^25^ for genes with valid gene imputation model estimated by at least one of these tools. The Manhattan plots of TWAS-O p-values of all six cell types are shown in **Fig. 2**.

**Figure 2.**
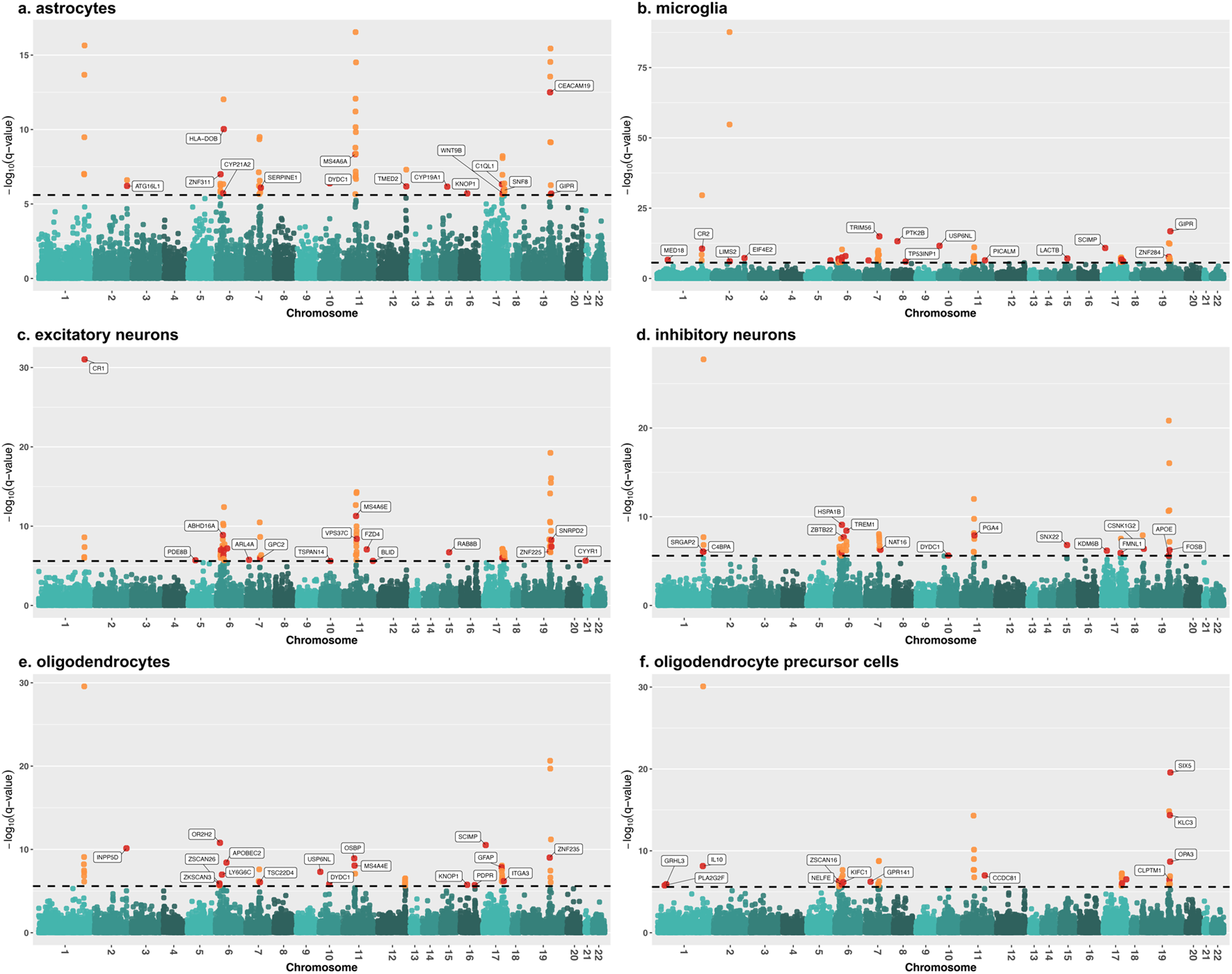
Manhattan plots (-log10(TWAS-O p-values)) by xWAS-O of AD dementia in astrocytes, microglia, excitatory neurons, inhibitory neurons, oligodendrocytes and oligodendrocyte precursor cells. Transcriptome-wide significant threshold -log10(2.5E-06) was plotted as the dashed horizontal line. Independent significant genes are colored in red and labelled, including the ones detected by GIFT and the ones having no overlapped test regions with other significant TWAS-O risk genes. Other non-independent TWAS-O risk genes are colored in orange.

Quantile-Quantile plots of TWAS results by these three individual tools and TWAS-O are shown in **Figs. S5-S10**. As expected, inflated false positive rates are observed in all TWAS results. This arises from several contributing factors: inflated GWAS summary statistics due to LD, overlapping test regions among nearby genes, and correlations among the genetically regulated expressions of neighboring genes. To account for the inflation, independent TWAS-O risk genes that either have fine-mapped GIFT p-values<0.05 or contain no overlapped test cis-genetic variants with other significant TWAS-O genes, are identified, colored in red, and labeled in the Manhattan plots (**Fig. 2**).

For astrocytes, we identified 65 significant TWAS-O risk genes, including 22 cell-type-specific ones (**Fig. 2A; Table S3**). Among these, 19 genes were previously linked to AD clinical or pathological traits in GWAS Catalog^37^ (**Table S3**), and 57 genes (88%) overlapped with previous bulk TWAS-O findings (**Table S1**)^20^. Using GIFT, we identified 15 independent significant risk genes (**Table 2; Fig. S12**) including 2 novel independent TWAS risk genes that have not been previously reported in GWAS nor bulk TWAS-O^20^ –– *CYP19A1* is involved in estrogen biosynthesis and previously reported to be associated with AD risk and neuroprotection^38^; *KNOP1* is linked to chromatin condensation and likely to contribute to AD pathology through transcription factor regulation^39^.

**Table 2:**
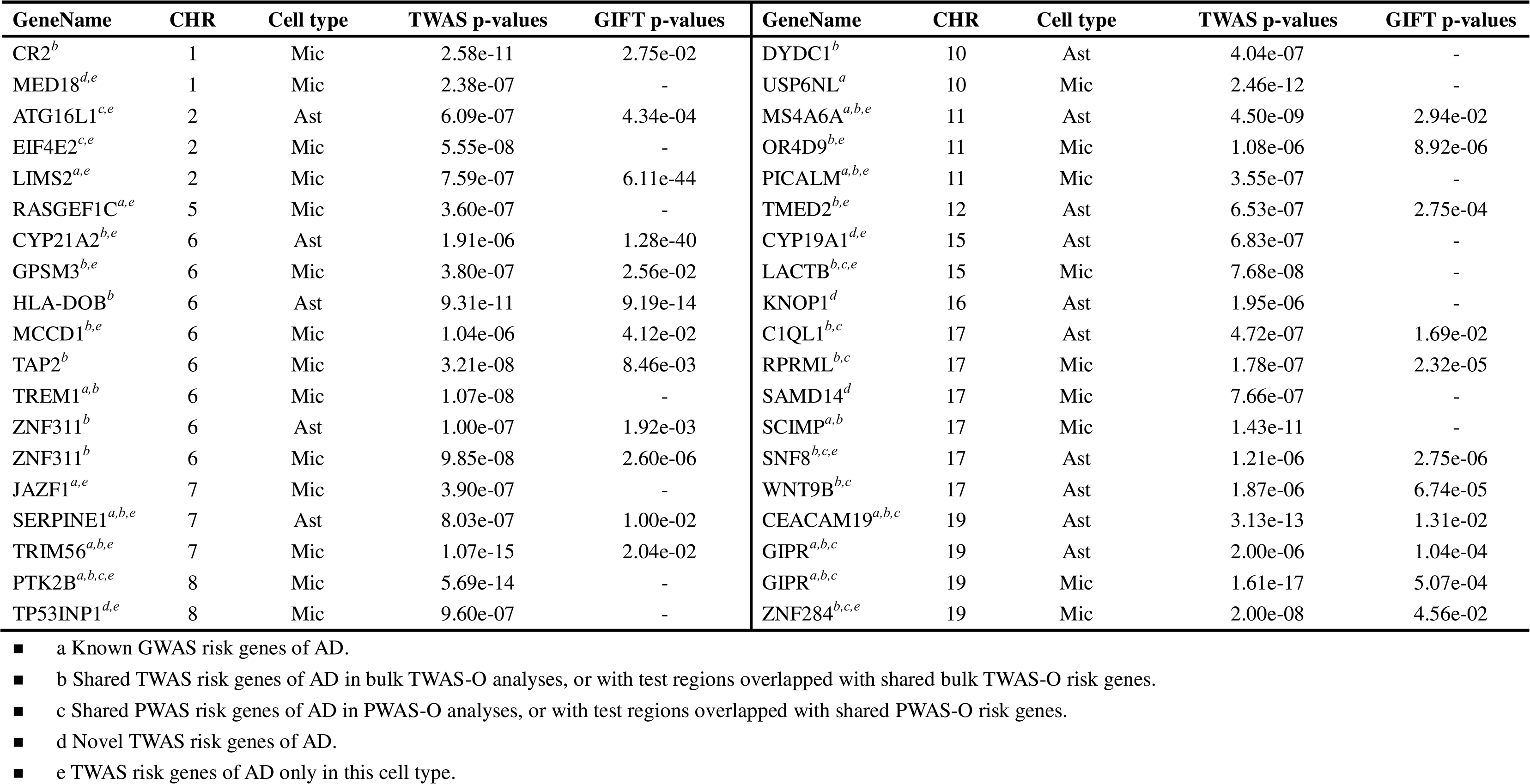
Independent significant cell-type-specific TWAS-O risk genes in astrocytes (Ast) and microglia (Mic). All significant genes here are either not overlapping with other genes, or independent significant genes identified by GIFT. NAs are shown as “-” in the table.

For microglia, we identified 64 TWAS-O significant risk genes, including 27 cell-type-specific ones (**Fig. 2B; Table S4**). Among these, 23 genes were previously reported to be associated with AD-related traits in the GWAS Catalog^37^ (**Table S4**), and 54 genes (84%) are overlapped with previous bulk TWAS-O findings (**Table S1**)^20^. Using GIFT, we identified 23 independent significant TWAS risk genes (**Table 2; Fig. S13**), including 2 novel findings (*MED18* and *TP53INP1*). *MED18* encodes a subunit of the Mediator complex, a key regulator of transcription, which has been implicated in neurodegenerative diseases^40^. Other related mediator subunits, such as *MED12* and *MED23*, that have been found to be deregulated in AD. This, suggests a potential role for *MED18* in transcriptional dysregulation and neuronal dysfunction in AD pathology^41^. *TP53INP1* is involved in oxidative stress responses, regulating p53 activity, that has been implicated in AD-related neurodegeneration^42^.

In neurons, we identified 77 significant TWAS-O risk genes in excitatory neurons and 49 in inhibitory neurons, including 35 and 21 cell-type-specific ones, respectively (**Tables S5-S6**). Among these, 24 and 13 genes were previously associated with AD-related traits in GWAS Catalog^37^; 69 (90%) and 48 (98%) genes overlapped with previous bulk TWAS-O findings^20^, respectively. Using GIFT, we identified 22 and 15 independent significant TWAS risk genes in excitatory and inhibitory neurons, respectively (**Table 3; Fig. S14-S15**). Two novel independent genes were identified in excitatory neurons (*PDE8B* and *BLID*). For example, *PDE8B*, which encodes a cAMP□specific phosphodiesterase, is enriched in neurons and microglia within cognition□critical regions such as the cerebral cortex and hippocampus. Additionally, its age□dependent up□regulation in 3×Tg□AD mice links impaired cAMP turnover to AD progression^43^. One novel independent gene was identified in inhibitory neurons: *KDM6B*, which encodes histone *H3K27* demethylase. This gene is essential for synaptic plasticity and cognitive function, and its Tau-dependent dysregulation has been linked to neuronal activity changes, cognitive deficits, and AD pathogenesis^44^.

**Table 3:** Independent significant cell-type-specific TWAS-O risk genes identified in excitatory neurons (Ex) and inhibitory neurons (In). All significant genes here are either not overlapping with other genes, or independent significant genes identified by GIFT. NAs are shown as “-” in the table.

| GeneName | CHR | Cell type | TWAS p-values | GIFT p-values | GeneName | CHR | Cell type | TWAS p-values | GIFT p-values |
| --- | --- | --- | --- | --- | --- | --- | --- | --- | --- |
| C4BPA<sup>b</sup> | 1 | In | 8.88e-07 | 8.04e-04 | BLID<sup>d,e</sup> | 11 | Ex | 2.38e-06 | - |
| CR1<sup>a,b</sup> | 1 | Ex | 9.25e-32 | 5.99e-03 | FZD4<sup>b,e</sup> | 11 | Ex | 8.49e-08 | - |
| SRGAP2<sup>b,e</sup> | 1 | In | 8.29e-07 | 8.60e-07 | MS4A6E<sup>a,b,e</sup> | 11 | Ex | 5.14e-12 | 1.34e-02 |
| PDE8B<sup>d,e</sup> | 5 | Ex | 1.97e-06 | - | PGA4<sup>b</sup> | 11 | In | 1.26e-08 | 1.40e-02 |
| ABHD16A<sup>b,e</sup> | 6 | Ex | 1.32e-09 | 9.39e-19 | VPS37C<sup>b,e</sup> | 11 | Ex | 3.93e-09 | 2.33e-02 |
| APOBEC2<sup>b</sup> | 6 | Ex | 6.30e-08 | - | RAB8B<sup>b,c,e</sup> | 15 | Ex | 1.96e-07 | - |
| HSPA1B<sup>b</sup> | 6 | Ex | 5.92e-08 | 4.03e-02 | SNX22<sup>b,c,e</sup> | 15 | In | 1.53e-07 | - |
| HSPA1B<sup>b</sup> | 6 | In | 8.15e-10 | 6.34e-03 | FMNL1<sup>b,c,e</sup> | 17 | In | 1.21e-06 | 2.59e-07 |
| LY6G6C<sup>b</sup> | 6 | In | 1.10e-06 | 1.24e-02 | HEXIM1<sup>b,c</sup> | 17 | Ex | 9.71e-07 | 1.16e-04 |
| NELFE<sup>b</sup> | 6 | Ex | 4.96e-07 | 3.48e-05 | KDM6B<sup>d,e</sup> | 17 | In | 6.62e-07 | - |
| TNXB<sup>a,b,e</sup> | 6 | Ex | 1.32e-07 | 3.32e-02 | WNT9B<sup>b,c</sup> | 17 | Ex | 9.36e-07 | 4.87e-22 |
| TREM1<sup>a,b</sup> | 6 | In | 3.76e-09 | 3.22e-03 | APOE<sup>a,b,c</sup> | 19 | In | 2.45e-06 | 2.91e-02 |
| ZBTB22<sup>b,e</sup> | 6 | In | 2.02e-08 | 7.62e-17 | CSNK1G2<sup>b,e</sup> | 19 | In | 4.08e-07 | 7.43e-43 |
| ZNF311<sup>b</sup> | 6 | Ex | 1.00e-07 | 1.02e-10 | FBXO46<sup>b,c,e</sup> | 19 | Ex | 3.76e-08 | 4.61e-02 |
| ARL4A<sup>c,e</sup> | 7 | Ex | 1.74e-06 | - | FOSB<sup>b,c</sup> | 19 | In | 5.76e-07 | 3.45e-02 |
| GPC2<sup>b</sup> | 7 | Ex | 1.32e-06 | 8.64e-05 | SNRPD2<sup>b,c,e</sup> | 19 | Ex | 5.51e-09 | 2.38e-03 |
| NAT16<sup>b,e</sup> | 7 | In | 5.29e-07 | 1.96e-03 | ZNF225<sup>a,b,c,e</sup> | 19 | Ex | 1.51e-07 | 1.37e-03 |
| DYDC1<sup>b</sup> | 10 | In | 2.31e-06 | - | CYYR1<sup>a,e</sup> | 21 | Ex | 2.24e-06 | - |
| TSPAN14<sup>a,b,e</sup> | 10 | Ex | 2.40e-06 | - |  |  |  |  |  |

- a Known GWAS risk genes of AD. - b Shared TWAS risk genes of AD in bulk TWAS-O analyses, or with test regions overlapped with shared bulk TWAS-O risk genes. - c Shared PWAS risk genes of AD in PWAS-O analyses, or with test regions overlapped with shared PWAS-O risk genes. - d Novel TWAS risk genes of AD. - e TWAS risk genes of AD only in this cell type.

In oligodendrocytes and Opcs, we identified 50 and 45 significant TWAS-O risk genes, including 20 and 16 cell-type-specific ones, respectively (**Tables S7-S8**). Among these, 18 and 14 genes were previously associated with AD-related traits in GWAS Catalog; 45 (90%) and 42 (93%) genes overlapped with previous bulk TWAS-O findings; GIFT identified 17 and 14 independent significant genes, respectively (**Table 4; Fig. S16-S17**). We identified three novel independent risk genes (*KNOP1*, *PDPR* and *ITGA3*) in oligodendrocytes, and two novel independent risk genes (*GRHL3* and *PLA2G2F*) in Opc.

**Table 4:** Independent significant cell-type-specific TWAS-O risk genes identified in oligodendrocytes (Oli) and oligodendrocyte precursor cells (Opc). All significant genes here are either not overlapping with other genes, or independent significant genes identified by GIFT. NAs are shown as “-” in the table.

| GeneName | CHR | Cell type | TWAS p-values | GIFT p-values | GeneName | CHR | Cell type | TWAS p-values | GIFT p-values |
| --- | --- | --- | --- | --- | --- | --- | --- | --- | --- |
| GRHL3<sup>d,e</sup> | 1 | Opc | 1.19e-06 | - | CCDC81<sup>b</sup> | 11 | Opc | 1.01e-07 | - |
| IL10<sup>b</sup> | 1 | Opc | 7.24e-09 | 3.18e-16 | MS4A4E<sup>a,b</sup> | 11 | Oli | 8.62e-09 | 7.10e-06 |
| PLA2G2F<sup>d,e</sup> | 1 | Opc | 1.66e-06 | - | OSBP<sup>b</sup> | 11 | Oli | 1.18e-09 | 1.23e-04 |
| INPP5D<sup>a,c,e</sup> | 2 | Oli | 7.26e-11 | - | KNOP1<sup>d</sup> | 16 | Oli | 1.74e-06 | - |
| APOBEC2<sup>b</sup> | 6 | Oli | 3.79e-09 | - | PDPR<sup>d,e</sup> | 16 | Oli | 1.88e-06 | - |
| KIFC1<sup>b,e</sup> | 6 | Opc | 6.65e-07 | 4.86e-08 | GFAP<sup>b,c,e</sup> | 17 | Oli | 1.37e-08 | 4.94e-02 |
| LY6G6C<sup>b</sup> | 6 | Oli | 1.05e-07 | - | ITGA3<sup>d</sup> | 17 | Oli | 6.21e-07 | - |
| NELFE<sup>b</sup> | 6 | Opc | 1.28e-06 | 4.08e-03 | LIMD2<sup>b,c,e</sup> | 17 | Opc | 3.02e-07 | - |
| OR2H2<sup>b,e</sup> | 6 | Oli | 1.58e-11 | 8.31e-04 | RPRML<sup>b,c</sup> | 17 | Opc | 9.07e-07 | 1.33e-05 |
| ZKSCAN3<sup>b</sup> | 6 | Oli | 1.70e-06 | 1.08e-06 | SCIMP<sup>a,b</sup> | 17 | Oli | 2.96e-11 | - |
| ZSCAN16<sup>b,e</sup> | 6 | Opc | 5.04e-07 | 5.48e-08 | CLPTM1<sup>a,b,c,e</sup> | 19 | Opc | 3.31e-07 | 1.64e-02 |
| ZSCAN26<sup>b</sup> | 6 | Oli | 1.22e-06 | 2.51e-02 | KLC3<sup>a,b,c</sup> | 19 | Opc | 4.33e-15 | 4.30e-03 |
| GPR141<sup>a,e</sup> | 7 | Opc | 5.91e-07 | - | OPA3<sup>a,b,c,e</sup> | 19 | Opc | 2.11e-09 | 2.56e-02 |
| TSC22D4<sup>b,e</sup> | 7 | Oli | 8.48e-07 | 8.53e-03 | SIX5<sup>b,c,e</sup> | 19 | Opc | 2.71e-20 | 5.54e-03 |
| DYDC1<sup>b</sup> | 10 | Oli | 1.48e-06 | - | ZNF235<sup>b,c,e</sup> | 19 | Oli | 9.52e-10 | 1.86e-10 |
| USP6NL<sup>a</sup> | 10 | Oli | 4.87e-08 | - |  |  |  |  |  |

- a Known GWAS risk genes of AD. - b Shared TWAS risk genes of AD in bulk TWAS-O analyses, or with test regions overlapped with shared bulk TWAS-O risk genes. - c Shared PWAS risk genes of AD in PWAS-O analyses, or with test regions overlapped with shared PWAS-O risk genes. - d Novel TWAS risk genes of AD. - e TWAS risk genes of AD only in this cell type.

### Comparison of PWAS-O and Cell-type-specific TWAS-O Findings

#### PWAS-O results

As shown in **Table 1**, 4069 valid protein abundance imputation models with CV *R*^2^ > 0.5% were obtained by TIGAR/DPR with median CV *R*^2^=0.013; 2003 valid protein abundance imputation models were obtained by PrediXcan/EN with median CV *R*^2^=0.041; 4710 valid protein abundance imputation models were obtained by FUSION/BestModel with median CV *R*^2^=0.0192. Scatter plots and histograms of the CV *R*^2^ of these three tools are presented in **Fig. S4**. Similarly, PWAS-O p-values were then derived from aggregating the PWAS p-values obtained from all three models by ACAT. Quantile-Quantile plots of PWAS-O and PWAS results by three tools are presented in **Fig. S11**.

We identified 21 PWAS-O significant risk genes of AD dementia with PWAS-O q-values<0.05, including 14 risk genes for AD-related traits previously listed in the GWAS Catalog^37^, 14 overlapped with previous bulk TWAS-O findings^20^, and 3 novel findings that were not associated with AD in previously reported GWAS/TWAS analyses (**Fig. 3, Table S2**). We fine-mapped 13 independent significant PWAS risk genes for AD dementia (**Fig. S18**).

**Figure 3.**
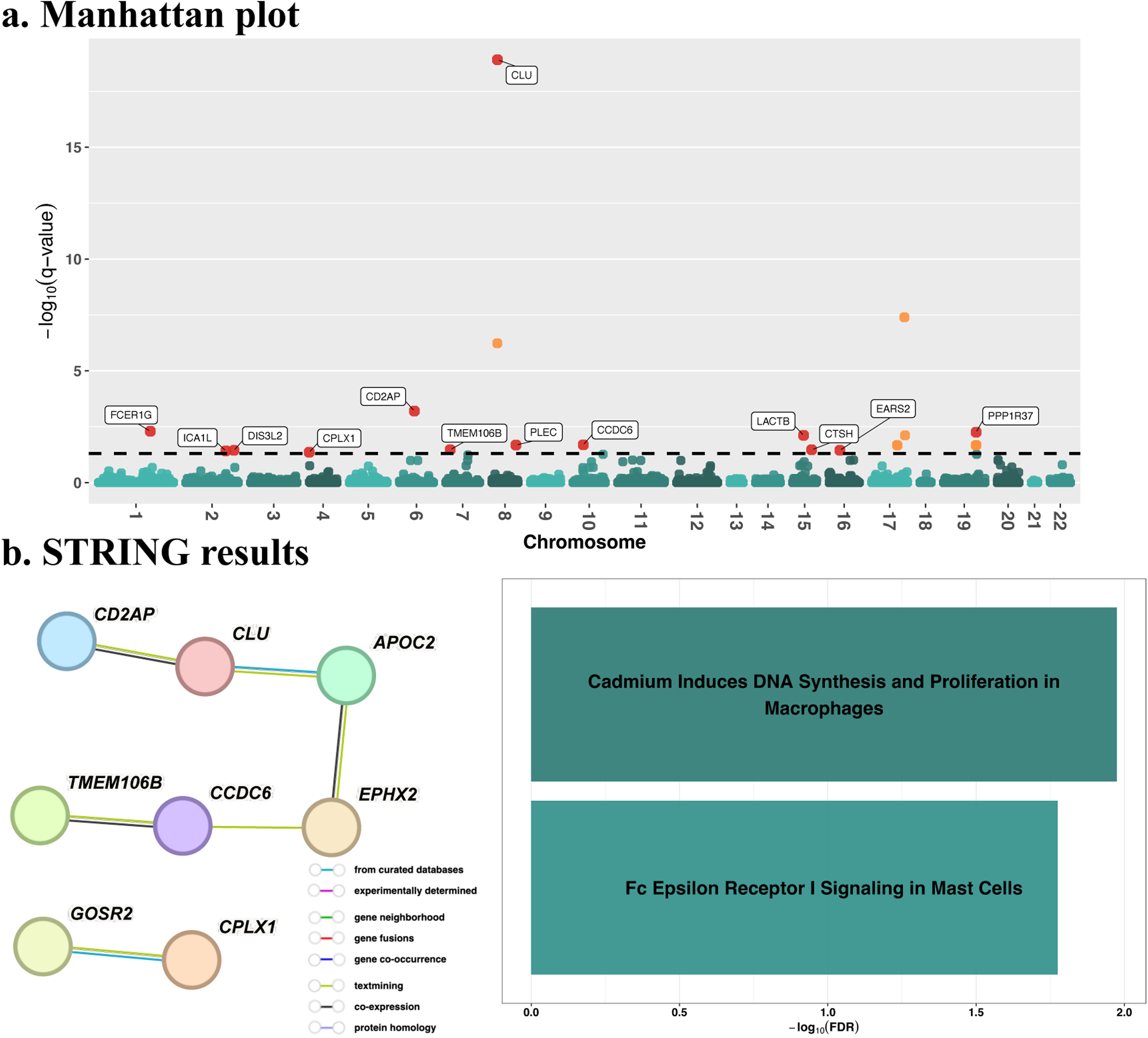
PWAS-O results of AD dementia. A.) Manhattan plot of -log10(PWAS-O q-values): Transcriptome-wide significance threshold -log10(0.05) was plotted as the dashed horizontal line. Independent significant PWAS-O risk genes are colored in red and labelled. Other non-independent ones are colored in orange. B.) PPI network and enriched phenotypes of significant PWAS-O risk genes: Edges in the PPI network plot represent physical PPI, with different colors representing different sources of connection evidence as shown in the legend. Phenotypes with known risk genes that are enriched with our PWAS-O findings are plotted, with the -log10(enrichment FDRs) in the x-axis.

#### Compare with cell-type-specific TWAS-O findings

To validate our findings, we compared the 91 detected independent significant cell-type-specific TWAS-O risk genes across all six brain cell types with the PWAS-O findings. Specifically, we checked if the test genetic regions of these independent genes overlapped with any protein coding genes that have PWAS-O q-values <0.05.

In astrocytes, we identified 6 out of 15 independent TWAS risk genes (40%) with test genetic regions overlapping with protein coding genes with adjusted PWAS-O q-values <0.05 (**Table 2**). Out of these 6 validated TWAS risk genes, we identified 4 genes (*C1QL1, SNF8, CEACAM19,* and *ATG16L1*) that are specific to astrocytes. *C1QL1*, a member of the C1q complement family, is implicated in Alzheimer’s disease and other neurodegenerative disorders due to its role in synapse formation, maintenance, and clearance of abnormal protein aggregates^45^*. CEACAM19*, a carcinoembryonic antigen-related cell adhesion molecule, that has been linked to mitochondrial pathology may play a role in the mechanisms underlying AD^46^. *ATG16L1*, encodes autophagy related 16 like 1 protein, whose activation has been studied to reduce Alzheimer’s disease-like pathology^47^.

In microglia, among 23 independent TWAS risk gene regions, we identified 6 (26%) overlapping with PWAS-O findings (**Table 2**). We identified 4 risk genes specific to microglia: *EIF4E2, PTK2B, LACTB,* and *ZNF284*. *EIF4E2*, encodes a cap□binding translation factor; its phosphorylation rises sharply in late□stage AD and tracks with hyper□phosphorylated Tau. These findings, link EIF4E2□mediated translation to neurofibrillary tangle formation^48^. *PTK2B*, a synaptic tyrosine kinase implicated in NMDA receptor regulation, dendritic spine plasticity, and tau phosphorylation pathways, has been shown to colocalize with both Aβ and tau pathology in the entorhinal cortex (EC) of AD. *PTK2B* expression has been found in microglia in the EC of AD, suggesting a potential role of neuroinflammatory mechanisms^49^. *LACTB*, whose reduced expression in myeloid cells—and its association with higher, AD□protective succinyl□carnitine—correlates with lower risk of Alzheimer’s disease^50^.

In neurons, we identified 7 of 22 independent TWAS risk gene regions in excitatory neurons (32%), and 3 of 15 in inhibitory neurons (20%), overlapping with PWAS-O findings (**Table 3**). We identified 7 genes specific to excitatory neurons: *ARL4A, RAB8B, HEXIM1, ZNF225, SNRPD2,* and *FBXO46*. *RAB8B* encodes a vesicle□trafficking Rab GTPase; experimental modulation of *RAB8B* activity eases AD□like pathology, marking it as a potential therapeutic target^51^. *HEXIM1*, encodes a P□TEFb□binding modulator that gates release of paused RNA polymerase□II, whose elevated expression in AD cortex perturbs activity□dependent transcription crucial for synaptic plasticity and memory^52^. *SNRPD2*, whose down□regulation in MCI and AD relaxes nuclear retention of lncRNAs/mRNAs, may hasten Alzheimer’s progression^53^. Moreover, we identified 4 genes: *SNX22*, *FMNL1*, and *FOSB*, specific to inhibitory neurons. Notably, previous research suggests that the dysfunction of *SNX22* may contribute to the development or progression of AD^54^.

In oligodendrocytes, we identified 3 of 17 independent TWAS risk gene regions (18%) overlapped with PWAS-O findings (**Table 4**), including 3 genes specific to oligodendrocytes (*INPP5D*, *ZNF235,* and *GFAP*). *INPP5D* encodes the microglial phosphatase□SHIP1, whose expression increases with amyloid burden and localizes to plaque□associated microglia in AD^55^. *GFAP* encodes glial fibrillary acidic protein; higher plasma *GFAP* mirrors tau accumulation in temporal cortex and predicts faster cognitive decline, underscoring astrocytic neuroinflammation in AD progression^56^.

In Opcs, we identified 6 of 14 independent TWAS risk gene regions (43%) overlapped with PWAS-O findings (**Table 4**), including 5 genes: specific to Opcs (*LIMD2*, *CLPTM1*, *KLC3*, *OPA3,* and *SIX5*). *LIMD2* has been identified as a hub gene for cell aging-immune/inflammation-related functions^57^, but its role in AD pathologies has not been reported. *CLPTM1*, a transmembrane protein, has been implicated in AD, with studies showing its association with both AD and epilepsy^39^. *SIX5*, a specific transcription factor (TF) plays a role in retinal function and is also involved in AD and age-related macular degeneration^58^.

We further compared significant PWAS-O and TWAS-O findings by calculating the pair-wise Jaccard Similarity Indexes^59^ using lists of unique names of significant genes. The Jaccard Similarity Index quantifies the overlaps of significant risk genes by PWAS-O, bulk TWAS-O, and cell-type-specific TWAS-O (**Fig. S19**). Here, only the same genes identified in both analyses are considered as overlapping. Higher Jaccard Similarity Index suggests higher percentage of overlapped risk genes. We found that cell-type-specific TWAS-O findings have 4%∼20% overlaps with each other^20^, with top overlaps found between astrocytes and excitatory neurons (20%), astrocytes and oligodendrocytes (17%), microglia and inhibitory neurons (16%).

### PPI Network and Enrichment Analyses

Using the STRING webtool^30^ (Methods), we conducted PPI network analyses using the lists of PWAS-O risk genes (**Fig. 3B**) and cell-type-specific TWAS-O risk genes of all six brain cell types (**Fig. 5**). The edges of the PPI network represent physical PPI and were colored according to different data sources as shown in the color legend. In addition to PPI network plots with protein coding genes that are connected with at least one other protein coding gene, FDR of pathways (obtained by pathDIP) enriched with our PWAS-O or cell-type-specific TWAS-O findings are shown by bar plots (**Fig. 3B**; **Fig. 5; Table S9**).

**Figure 4.**
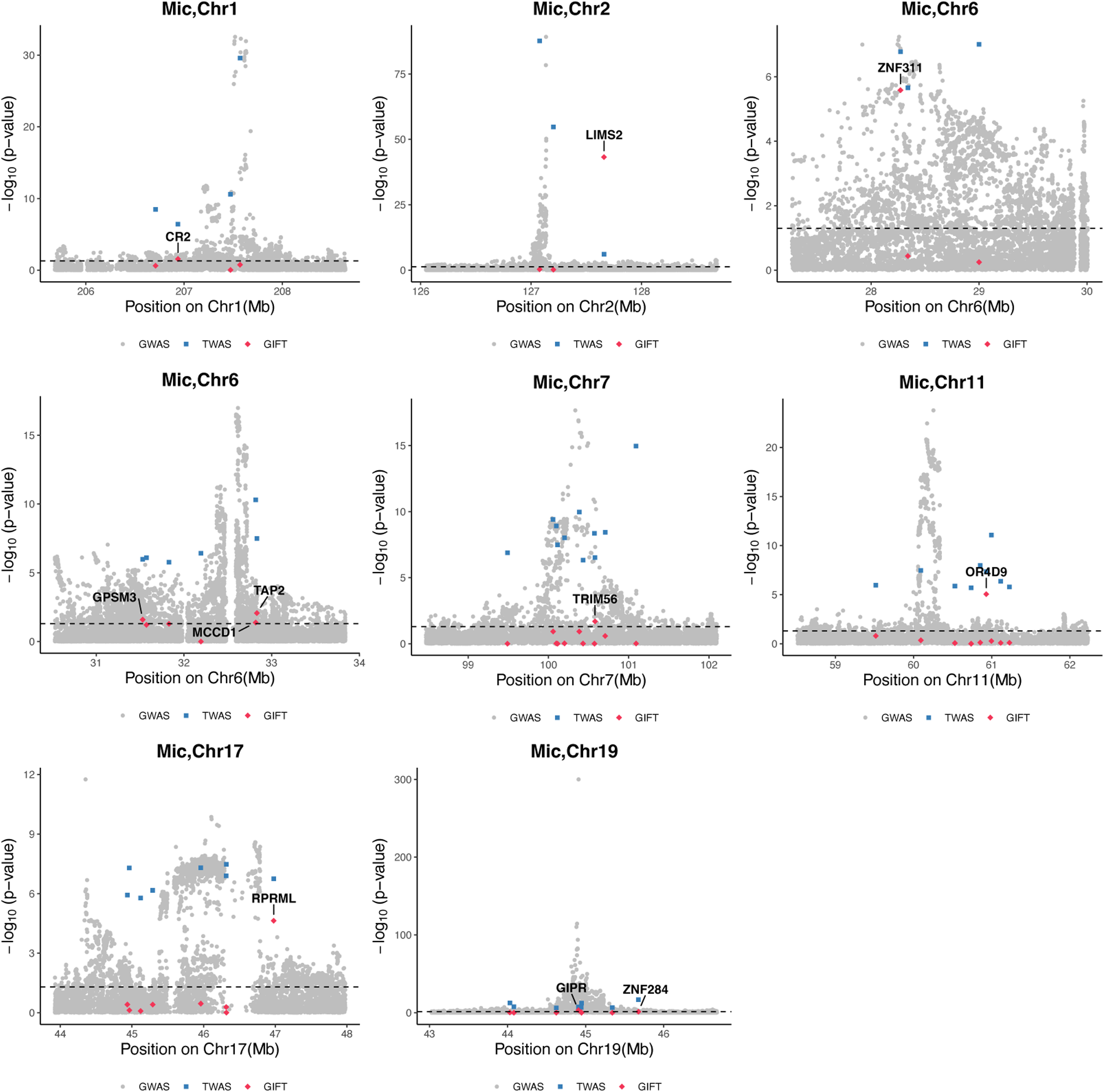
Fine mapping results of TWAS-O results of AD dementia by GIFT in microglia. The -log10(p-values) were plotted on the y-axis, and -log10(0.05) was plotted as the dashed horizontal line. Gray dots: SNPs by GWAS; Blue squares: genes identified by TWAS; Red diamond: results by GIFT. Causal significant genes are labeled.

**Figure 5.**
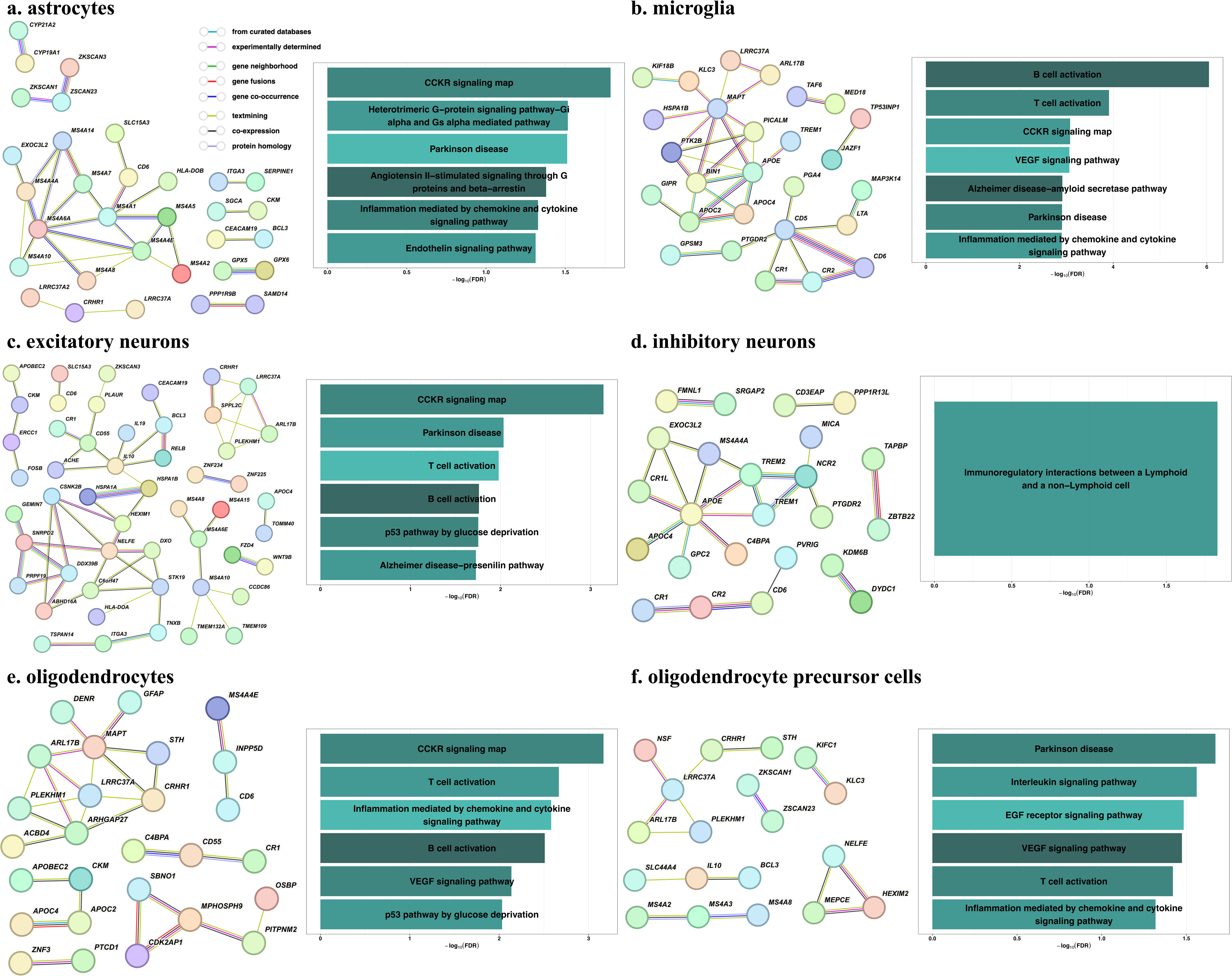
PPI network and enrichment analyses results with cell-type-specific TWAS-O risk genes of AD dementia in six major brain cell types. Edges represent physical PPI, with different colors representing different sources of connection evidence. PWAS risk genes not connected in the network are not included in the network plot.

Eight of 21 (38.1%) PWAS-O risk genes were interconnected in the PPI network plot (**Fig. 3B**). *APOC2* is also identified as a TWAS-O risk gene in microglia and oligodendrocytes, which encodes a protective apoE isoform that lowers AD risk via both Aβ□dependent and independent pathways and is associated with increased longevity^60^. Two biological processes emerged as significantly enriched with PWAS-O risk genes (Benjamini–Hochberg FDR□<□0.05): The biological process of “Cadmium induces DNA synthesis and proliferation in macrophages” (FDR=1.06E-02) is enriched with *CLU*, *DIS3L2,* and *PPP1R37*. *CLU* encodes a secretory protein predominantly synthesized in brain astrocytes that are strongly induced by neurons sensitive to AD risk factors, that would inhibit Aβ degradation and promote Aβ generation^61^. *PPP1R37* encodes protein phosphatase 1 regulatory subunit 37, which has been reported to be associated with AD^62^. The other biological process of “Fc Epsilon Receptor I Signaling in Mast Cells” (FDR = 1.68E-02) is enriched with *CD2AP*, *FCER1G* and *PPP1R37*, and Mast Cells were associated with AD progression^63^.

Across all six brain cell types, 42.2%-66.2% of our detected cell-type-specific TWAS-O risk genes formed interconnected networks, featuring well-established AD risk genes such as *APOE*^22^, *BIN1*^23^, and *MAPT*^24^ as hubs of the PPI networks. Importantly, *APOE*, encodes apolipoprotein□E, the brain’s chief lipid□transport protein; its ε4 isoform accelerates and amplifies amyloid□β plaque seeding, making APOE the principal genetic driver of Alzheimer’s risk^22^.

In astrocytes, 32 of 65 (49.2%) TWAS-significant genes formed PPI networks (**Fig. 5A**), with known AD risk genes *MS4A1* and *MS4A6A* as hub nodes. *MS4A1*, which is known to form or serve as a functional component of a Ca2+-permeable cation channel, has been shown involved in AD pathogenesis^64^. *MS4A6A* is linked to cerebrospinal fluid soluble TREM2 levels regulation and associated with AD progression^64,65^. Specifically, *MS4A1*, *HEXIM1*, *ATG16L1* and *PFKFB2* were enriched in Parkinson disease pathway (FDR = 3.08E-02), which has neuropathological overlap with AD^66^. The recurrent inclusion of *BCL3* and *PPP1R9B* across multiple pathways underscores a pro□inflammatory, GPCR□driven astrocytic response that could modulate APP processing and neurovascular tone in AD^67,68^.

In microglia, 27 of 64 (42.2%) TWAS-O risk genes were interconnected in PPI networks (**Fig. 5B**), with known AD risk genes *CD5*, *MAPT*, *BIN1*, and *APOE* as hubs in the PPI networks. Importantly, *BIN1*, *MAPT,* and *MAPKAPK2*, among others, were enriched in B-cell activation (FDR=8.99E-07) and CCKR signaling map (FDR=8.33E-04), reinforcing the role of microglial immune response in AD. *BIN1*, with *BIN1*-dependent alterations in neuronal properties, has been associated with AD pathophysiology^23^. *MAPT* encodes the predominant component of neurofibrillary tangles tau, which are neuropathological biomarker of AD pathology^24^. Genes such as *CD5*, *MAPKAPK2, MS4A2*, and *PTK2B* were associated with T-cell activation (FDR=1.23E-04), where T cells are often dysfunctional in Alzheimer’s disease and are involved in AD pathology^69^. Additionally, several risk genes, including *CD5*, *MAPKAPK2, MAPT, and PTK2B* were also enriched in VEGF signaling (FDR=8.64E-04), a known pathway implicated in AD progression^70^. Interestingly, the convergence of B□ and T□cell activation, VEGF, and cytokine pathways suggests that microglial immune signaling and angiogenic cross□talk form an integrated axis driving AD□related neuroinflammation.

In neurons, 51 of 77 (66.2%) excitatory neuron genes were interconnected in PPI networks (**Fig. 5C**), with known AD risk genes *SPPL2C, MS4A6E,* and *IL10*^71^ as hubs in the PPI networks. Enrichment analysis revealed that excitatory neuron risk genes were associated with CCKR signaling map (FDR=7.20E-04) and AD-presenilin pathway (FDR=1.88E-02), the most common cause of familial Alzheimer’s disease^72^.

In inhibitory neuron results, 24 of 49 (49.0%) genes were interconnected in PPI networks (**Fig. 5D**), with known AD risk genes *APOE* and *TREM2* as hubs in the PPI networks. Particularly, inhibitory neuron risk genes were enriched in immunoregulatory interactions between lymphoid and non□lymphoid cells (FDR=1.48E-02), underscoring a selective involvement of inhibitory neuronal ligands (TREM1/TREM2) in neuron–immune crosstalk.

In oligodendrocytes, 27 of 50 (54.0%) formed PPI networks (**Fig. 5E**), with known AD risk genes *MAPT* and *APOC2* as hubs in the PPI networks. Oligodendrocyte risk genes were significantly enriched in CCKR signaling map (FDR=6.81E-04) and T cell activation (FDR=2.15E-03). Genes shared between oligodendrocytes and microglia (*MAPKAPK2*, *MAPT*) were enriched in B cell activation (FDR=3.09E-03), emphasizing the importance of immune cells activation in AD progression.

In oligodendrocyte precursor cells, 19 of 45 (42.2%) risk genes formed PPI networks (**Fig. 5F**), with known AD risk genes *LRRC37A*^73^ and *IL10*^71^ as hubs in the PPI networks. Risk genes in oligodendrocyte precursor cells were also enriched in VEGF signaling pathway (FDR=3.36E-02) and T cell activation (FDR=3.8E-02). Interestingly, *BCL3*, *IL10,* and *MS4A2* were enriched in Interleukin signaling pathway (FDR=2.73E-02), that is a key inflammatory pathway associated with AD pathology^74^.

Several pathways were significant in multiple cell types, e.g., CCKR signaling map, T□cell activation, B□cell activation, VEGF signaling pathway, inflammation mediated by chemokine and cytokine signaling pathway, and Parkinson disease. These findings suggest that similar immune, vascular, and neurodegeneration processes affect many cell types in AD. We also identified cell-type-specific significant pathways. For example, “endothelin signaling pathway” associated with astrocytes in AD pathogenesis^75^, and “heterotrimeric G-protein signaling pathway-Gi alpha and Gs alpha mediated pathway”. G-protein receptor was implicated in AD pathology^76^, was only identified in astrocytes. “Alzheimer disease□amyloid secretase pathway”, associated with AD related amyloid β-peptide^77^, was uniquely identified in microglia.

## Discussion

Leveraging multiple complimentary statistical methods^5–8^ by xWAS-O, we identified 223 cell-type-specific significant risk genes with TWAS-O p-values <2.5E-06, which were further fine-mapped to 91 independent genes by GIFT^21^ including 11 novel findings. Compared to previous cell-type-specific TWAS analyses by FUSION^8^ using the same snRNA-seq and GWAS summary data^19^, we detected more significant TWAS risk genes using the more stringent genome-wide significance threshold of p-values<2.5E-06 (vs. FDR<0.05), 223 vs. 165 with Jaccard Index 9% in the same 6 major brain cell types. Also, 40 out of our 91 independent cell-type-specific TWAS-O findings are shown overlapped with findings by FUSION in this previous study.

Another novel perspective of this study is that we conducted analogous PWAS-O analyses using large-scale proteomics data^78,79^ of DLPFC in the same ROS/MAP cohorts and the same GWAS summary data^4^. We identified 21 significant AD risk genes including 13 fine-mapped independent ones. Among these 21 PWAS-O risk genes, 14 were overlapped with previous PWAS findings^18,80^ using a subset of the ROS/MAP proteomics data (n=355) and different GWAS summary data of AD dementia^81,82^. Comparing our cell-type-specific TWAS-O findings to these PWAS-O findings, we validated 29 out of 91 (31.9%) independent cell-type-specific TWAS-O risk genes, which were overlapped with at least one of these 21 PWAS-O significant genes.

Further PPI network analyses of lists of our cell-type-specific TWAS-O and PWAS-O risk genes detected inter-connected PPI networks with known AD risk genes, such as *SPPL2C*^83^, *APOE*^22^, *BIN1*^23^, and *MAPT*^24^, as hub genes. Our detected cell-type-specific TWAS-O risk genes were found enriched in several shared AD-related pathways, such as CCKR signaling map, B cell activation, T cell activation, and p53 pathway by glucose deprivation (**Fig. 5**). Cell-type specific enriched pathways were also detected, such as Endothelin signaling pathway and Angiotensin II-stimulated signaling through G proteins and beta-arrestin for astrocytes, and Alzheimer disease□amyloid secretase pathway for microglia, and Alzheimer disease-presenilin pathway for excitatory neurons. Our detected PWAS-O risk genes were found enriched with known risk genes of Cadmium Induces DNA Synthesis and Proliferation in Macrophages and Fc□Epsilon Receptor□I Signaling in Mast Cells (**Fig. 3B**). These results suggest that important cell-type-specific TWAS risk genes could help illustrate cell-type-specific mechanisms underlying AD, and that the PWAS analyses would complement TWAS analyses by identifying post-transcriptionally regulated risk genes that are enriched in lipoprotein measurements.

Especially, our results reinforce the crucial role of microglia and astrocytes in AD. TWAS-O analysis in microglia showed the highest number of independent risk genes (23), and 27 out of 64 (42.2%) TWAS-O risk genes in microglia exhibited robust PPI connectivity. Well-established AD genes like *APOE*^22^, *MAPT*^24^, and *BIN1*^23^ served as key hubs in the PPI networks. Genes such as *CD5*^84^ and *MAPKAPK2*^85^ were shown enrichment in T cell activation and VEGF signaling, supporting microglial involvement in neuroinflammation and cognitive aging. Importantly, novel microglia-specific risk genes (*MED18*^40,41^ and *TP53INP1*^42^) were identified, suggesting transcriptional regulation and oxidative stress pathways may be underappreciated contributors to microglial-mediated AD risk.

In astrocytes, 32 out of 65 (49.2%) TWAS-O genes were interconnected in PPI networks, with hubs genes *MS4A1*^64^ and *MS4A6A*^65^ that were previously linked to amyloid processing and mitochondrial function. The discovery of novel astrocyte-specific risk genes—including *CYP19A1*^38^ and *KNOP1*^39^—highlights emerging mechanisms involving estrogen synthesis, and chromatin condensation, which may intersect with astrocyte-mediated neuroinflammatory and homeostatic functions. In both neuronal and oligodendrocyte lineage cells, over 40% of TWAS-O risk genes formed interconnected PPI networks, highlighting coordinated molecular mechanisms relevant to AD, suggesting strong functional organization within each cell type.

Despite these important findings of cell-type-specific TWAS-O and PWAS-O risk genes of AD in this study, several limitations should be acknowledged. First, inflation is observed in our cell-type-specific TWAS-O and PWAS-O results. This is expected due to the inflation in GWAS summary data due to LD, overlapped test cis-xQTL of nearby genes, and correlated genetically regulated gene expression or protein abundances of nearby genes. We expect to calibrate the false positive inflation rates by fine-mapping these results for independent signals, as implemented in this study. Second, we only studied cis-xQTL in this study, which might miss risk genes whose gene expression or protein abundances are affected by trans-xQTL. Further studies are ongoing about exploring both cis- and trans-xQTL in xWAS analyses of AD dementia. Third, although 31.9% of cell-type-specific TWAS-O risk genes were also validated in the PWAS-O analyses, follow-up biological validations are needed for understanding the underlying biological mechanisms of these detected risk genes.

In summary, we demonstrated that by leveraging large-scale snRNA-seq and proteomics data of DLPFC, xWAS-O findings provide a more granular characterization of AD risk genes. In particular, by leveraging multiple complementary statistical method as implemented in three tools of TIGAR^5,6^, PrediXcan^7^, and FUSION^8^, our analyses show higher power than using only one tool. We further provided a list of 91 fine-mapped cell-type-specific TWAS risk genes (11 novel) and 13 fine-mapped PWAS risk genes of AD dementia. From a translational perspective, our results provide a prioritized list of cell-type-specific candidate TWAS risk genes for functional validation and therapeutic targeting, 31.9% of which were validated by PWAS-O analyses. Several of the identified genes were shown having established interactions with known AD risk genes (e.g., *APOE*, *BIN1*, *MAPT*) in PPI networks, suggesting that they may contribute to shared pathogenic pathways. Our findings emphasize that risk genes of AD dementia might have genetic effects mediated through transcriptomic data of different cell types, and that our detected cell-type-specific TWAS risk genes provides valuable insights into disease etiology of AD. Our findings in this study underscore the importance of cell-type-specific TWAS and PWAS analyses in unraveling the molecular mechanisms underlying AD.

## Methods

### xWAS-O analytical framework

As shown in our recently published PWAS-O paper^18^, imputation models for gene expression quantitative traits, here we used cell-type-specific expression, or protein abundance quantitative traits (i.e., molecular trait ***T_g_***), are first trained in Stage I using individual-level genetic and transcriptomic/proteomic data from reference panels. Cis-genotype data (***G***, ±1 Mb of the transcription start and termination sites of the target gene) are used as predictors in the assumed multiple linear regression model (***Equation 1***), with effect sizes (***W***) estimated by multiple complementary statistical methods.

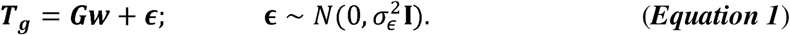

Covariates such as age, sex, top 20 Probabilistic Estimation of Expression Residuals (PEER) factors^86^, top 5 genotype principal components (PC), and batch effects were regressed out from the raw log2 transformed gene expression (or protein abundance) quantitative traits. Residuals after adjusting for these covariates are taken as response variables ***T_g_***.

xWAS-O employs three imputation models: (i) nonparametric Bayesian Dirichlet Process Regression (DPR) as implemented by TIGAR tool^5,6^, (ii) penalized regression with Elastic-Net penalty (initially proposed in the PrediXcan^7^ tool, also implemented in the TIGAR tool), (iii) best model selected by FUSION tool^8^ according to CV *R*^2^, out of single best eQTL (Top-QTL), penalized regression with LASSO penalty, penalized regression with Elastic-Net penalty, best linear unbiased predictor computed from all SNPs (BLUP). For each quantitative trait of a test gene, three sets of xQTL weights (***ŵ***) will be estimated by TIGAR/DPR, PrediXcan/EN, and FUSION/BestModel.

In Stage II, xWAS-O first uses these three sets of xQTL weights (***ŵ***) from Stage I as variant weights (***Equation 2***) to implement the burden test to compute a gene-based association Z-score test statistic, with respect to the phenotype in the summary-level GWAS test data^87^. The gene-based association Z-score test statistic is derived by the S-PrediXcan paper^87^, given by

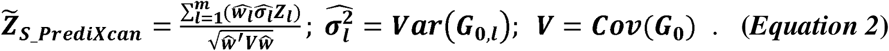

Here, ***Z_i_*** denotes the single-variant Z-score test statistics in GWAS summary data for the *l^th^* genetic variant, 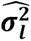 denotes genotype variance of the *l^th^* genetic variant, and ***V*** denotes the genotype variance-covariance matrix. Both 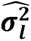 and ***V*** could be approximated from individual-level genotype data of an external reference panel of the same ancestry as the test GWAS data (with genotype matrix ***G_O_***). Reference LD derived from the WGS genotype data of the ROS/MAP cohorts^26^ was used for deriving xWAS Z-score test statistics in this study. Gene-based association test p-values can be easily derived from the Z-score statistic in ***Equation 2*** that has a standard normal distribution under the null hypothesis of no association exists between genetically regulated molecular traits and the phenotype of interest.

To enhance the robustness and statistical power of gene-based association testing, xWAS-O utilizes the Aggregated Cauchy Association Test^25^ (ACAT) to combine p-values from all three imputation tools as:

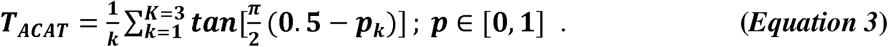

Here, ***K*** denotes the total number of xWAS p-values per gene, and ***p_k_*** denotes the p-value obtained from a gene-based association test by each of these three tools. ACAT generates an omnibus test p-value for each test molecular trait of a test gene by aggregating p-values derived from three complementary statistical models, which is then used to identify significant xWAS risk genes^18^. By comprehensive simulation studies and real studies of complex traits, previous studies^18,88^ have shown that improved power and calibrated type I error rates were obtained using ACAT omnibus xWAS p-values.

### Fine-map xWAS-O risk genes by GIFT

Gene-based Integrative Fine-mapping through conditional TWAS^21^ (GIFT) was implemented to fine map xWAS-O risk genes. First, we extended each gene’s start and end positions by 1Mb, thereby defining the flanking regions. Any overlapping flanking regions among xWAS-O risk genes were then merged into a single genomic region. Second, for each genomic region, GIFT jointly models genetically regulated gene expression or protein abundance components of all genes in the target genomic region, while explicitly accounting for correlations among the genetically regulated gene molecular traits and LD among cis-SNPs. As a result, GIFT estimates causal effects for all test genes in the target genomic region, effectively pinpointing putative causal genes. When individual-level data are unavailable, GIFT leverages summary statistics and LD reference panels to model gene expression correlation and cis-SNP LD, enabling robust fine-mapping of candidate causal genes.

For each gene i, the marginal association Z-scores from the xQTL data are modeled as:

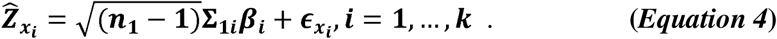

Here, *n*_1_ is the sample size for xQTL summary data, Σ_1*i*_ is the LD correlation matrix of cis-SNPs for gene *i* in the xQTL summary data, *β_i_* represents cis-SNP effect sizes on gene expression or protein abundance, and *ϵ_xi_* captures residual errors.

After that, the GWAS summary statistics for all cis-SNPs in the genomic region are modeled as:

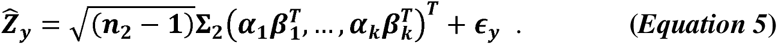

Here, *n_2_* is the sample size for GWAS summary data, Σ_2_ is the LD correlation matrix of all cis-SNPs across genes in the GWAS summary data, *β_i_* are from ***Equation 4***, *α_i_* denotes the conditional effect of the genetically regulated gene expression (GReX) or protein abundance (GRP) of gene *i* on the phenotype, and *ϵ_y_* represents residual errors. This equation jointly tests the association of each gene’s GReX or GRP with the phenotype while conditioning on the effects of other genes in the region.

Since our omnibus framework won’t have a combined weight for all three model, we choose to implement GIFT with summary statistics. We used the GWAS summary data of AD dementia^4^, xQTL summary data generated by BGW-TWAS tool^89^ using individual level xQTL data, along with the reference LD (i.e., genotype correlation matrix) derived from the individual-level genotype data of both UK Biobank^90^ and ROSMAP^26^ reference panels. Reference LD derived from different reference panels are recommended for matching the same ancestry as the GWAS and xQTL data to better estimated the positive semidefinite matrices in the ***Equation 4 and 5***.

### PPI network analyses by STRING

The STRING^30–32^ webtool integrates public data sources of protein interaction and analyzes the protein-protein interaction (PPI) network connectivity of proteins. Protein-protein edges represent the functional association, colored with six different connections –– curated databases, experiments, text mining, co-expression, gene co-occurrence and protein homology. Gene co-occurrence association predictions are based on whole-genome comparisons. We utilized the STRING webtool to conduct PPI network analyses with the list of cell-type-specific TWAS-O and PWAS-O risk genes.

### Enrichment analyses pathDIP

PathDIP□5^33^ integrates pathways from 6,500□plus curated records across many databases, unifying them with a KEGG□/Reactome□based ontology so enrichment results can be filtered, consolidated, and mapped into 53 functional categories, with FDR q-value by both BH-method and Bonferroni. We can pick specific pathway sources or types, analyze genes or metabolites, and explore drug□gene networks via Drugst.One. Enrichment analysis aims to detect pathways that are significantly enriched with xWAS-O risk genes verses random genes. We utilized the pathDIP webtool to conduct enrichment analyses with respect to data bases of Panther^34^, PathBank^35^ and Reactome^36^ with the list of cell-type-specific TWAS-O and PWAS-O risk genes.

### ROS/MAP omics data

The ROS/MAP genetics, snRNA-seq, and proteomics data originate from two longitudinal, community-based clinic pathological cohort studies (ROS and MAP)^26^. Participants were recruited without known dementia at the time of enrollment and agreed to annual clinical evaluations and brain donation at the time of death. Each study received approval from the Institutional Review Board of Rush University Medical Center, and at the time of enrollment all participants sign informed consent forms for the studies, Anatomical Gift Act agreements, and repository consents^91^.

In this study, snRNA-seq data were profiled from 436 postmortem specimens from dorsal lateral prefrontal cortex (DLPFC) from ROS/MAP decedents, yielding transcriptomic profiles of 1,546,794 cells^19^. Our analyses focused on six major brain cell types: astrocytes (Ast), microglia (Mic), excitatory neurons (Ex), inhibitory neurons (In), oligodendrocytes (Oli), and oligodendrocyte precursor cells (Opc). For each gene, its pseudo-bulk expression quantitative trait was generated for each cell type, by summarizing the raw reads counts across all cells of the same cell type. The raw read counts in single nuclei were combined into pseudo-bulk RNA-seq counts per gene per cell type, which were then converted to counts per million (CPM) and log2 transformed for improved normality.

Human brain proteomic profiles were quantified by tandem mass spectrometry from DLPFC of 716 postmortem brains donated by participants of European ancestry from the ROS/MAP cohorts^78,79^. As described in a previous publication^92^, protein abundances were normalized by scaling each protein abundance with respect to a sample-specific total protein abundance, and proteins with missing values in more than 50% of the participants were excluded from the analyses. Protein abundance ratios related to baseline (sample-specific total protein abundance) were log2 transformed for improved normality, referred to as protein abundance traits, with missing values imputed by the corresponding mean in the cohort. Outlier samples were removed using the iterative principal component analysis (PCA) method as describe in previous publication of PWAS analyses^92^.

Confounding covariates were regressed out from the cell-type-specific pseudo-bulk expression quantitative traits and protein abundance traits. For expression quantitative traits, sex, age at death, study cohort (ROS or MAP), and top 20 PEER^86^ factors were regressed out. For protein abundance traits, sex, age at death, postmortem interval, batches (i.e., sequencing batch for gene expressions or MS2 versus MS3 mass spectrometry reporter quantification mode for protein abundances), and top 5 genotype principal components were regressed out.

Genotype data of ROS/MAP samples were profiled by whole genome sequencing (WGS)^27^. Genetic variants with Hardy-Weinberg equilibrium p-values <10^-s^ and minor allele frequency >1% were used for estimating cis-xQTL weights.

### GWAS summary data of AD dementia by Bellenguez et. al. 2022

The most recent GWAS summary data (n=∼789K) of AD dementia^4^ were generated by meta-analysis, including clinically diagnosed AD cases and proxy AD and related dementia (proxy-ADD), resulting in a total of approximately 64,498 clinically diagnosed AD cases, 46,828 proxy-ADD cases, and 677,663 controls.

## Data Availability

All omics data of ROS/MAP samples are available from Synapse (https://www.synapse.org/Synapse:syn3219045). ROS/MAP resources by Bennett et. al. 2018 can be requested at www.radc.rush.edu and www.synapse.org. GWAS summary data by the Bellenguez et. al. 2022 are available from European Bioinformatics Institute GWAS Catalog (https://www.ebi.ac.uk/gwas/) under accession no. GCST90027158. Trained cell-type-specific cis-eQTL weights, cis-pQTL weights, and TWAS/PWAS summary data will be shared through SYNAPSE once this work is published.

## Code Availability

The analysis code used to produce all results in this work is available at https://github.com/Leo-LiuQiang/CTS-TWAS.

## Ethics declarations

ROS/MAP studies received approval from an Institutional Review Board of Rush University Medical Center, with all participants signing informed consent forms, Anatomical Gift Act agreements, and repository consents. All individual-level snRNAseq and proteomics data analyzed in this work were de-identified.

## Acknowledgements

This work is supported by the National Institutes of Health (NIH), National Institute of General Medical Sciences (NIGMS, R35GM138313, for Q.L. and J.Y), ROSMAP is supported by P30AG10161, P30AG72975, R01AG17917. R01 AG015819, U01 AG072572, and U01 AG046152.

## Supplementary Information

19 supplementary figures and 9 supplementary tables in the supplementary excel are provided.

