## Supplementary Figures for "Cell-type-specific Transcriptomic-wide Association Studies Detected 91 Independent Risk Genes for Alzheimer’s Disease Dementia"

**Fig. S1. Pairwise comparison of CV $R^2$ of cell-type-specific gene expression imputation models by TIGAR/DPR, PrediXcan/EN, and FUSION/BestModel in astrocytes and microglia.**

Each panel presents scatter plots comparing CV $R^2$ between pair-wise models, with each dot representing a gene. Colors indicate different gene groups: genes with CV $R^2$ > 0.5% in both compared models (Both:teal), in only one of the two compared models (TIGAR-only:red, Elastic Net-only:green, or FUSION-only:blue), or in neither models (Neither:gray).

**A. Astrocytes**

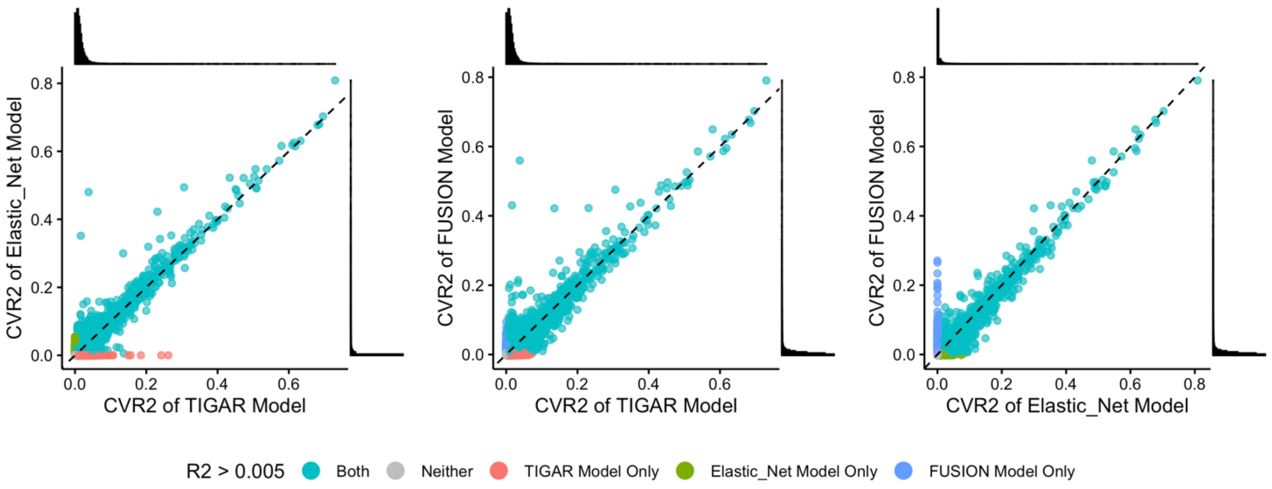


**B. Microglia**

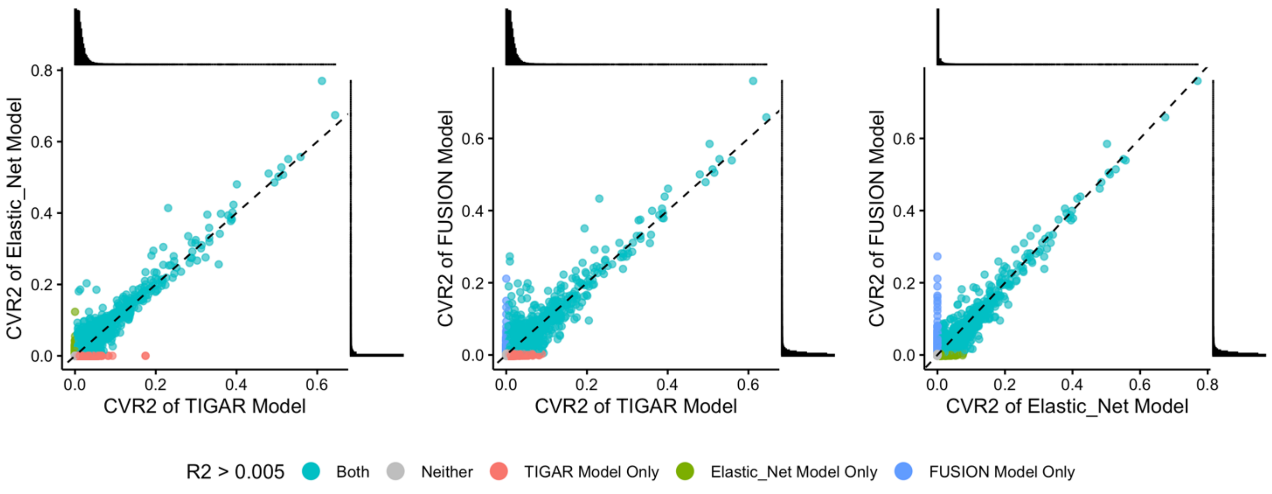

**Fig. S2. Pairwise comparison of CV $R^2$ of cell-type-specific gene expression imputation models by TIGAR/DPR, PrediXcan/EN, and FUSION/BestModel in Excitatory Neurons and Inhibitory Neurons.**

Each panel presents scatter plots comparing CV $R^2$ between pair-wise models, with each dot representing a gene. Colors indicate different gene groups: genes with CV $R^2$ > 0.5% in both compared models (Both:teal), in only one of the two compared models (TIGAR-only:red, Elastic Net-only:green, or FUSION-only:blue), or in neither models (Neither:gray).

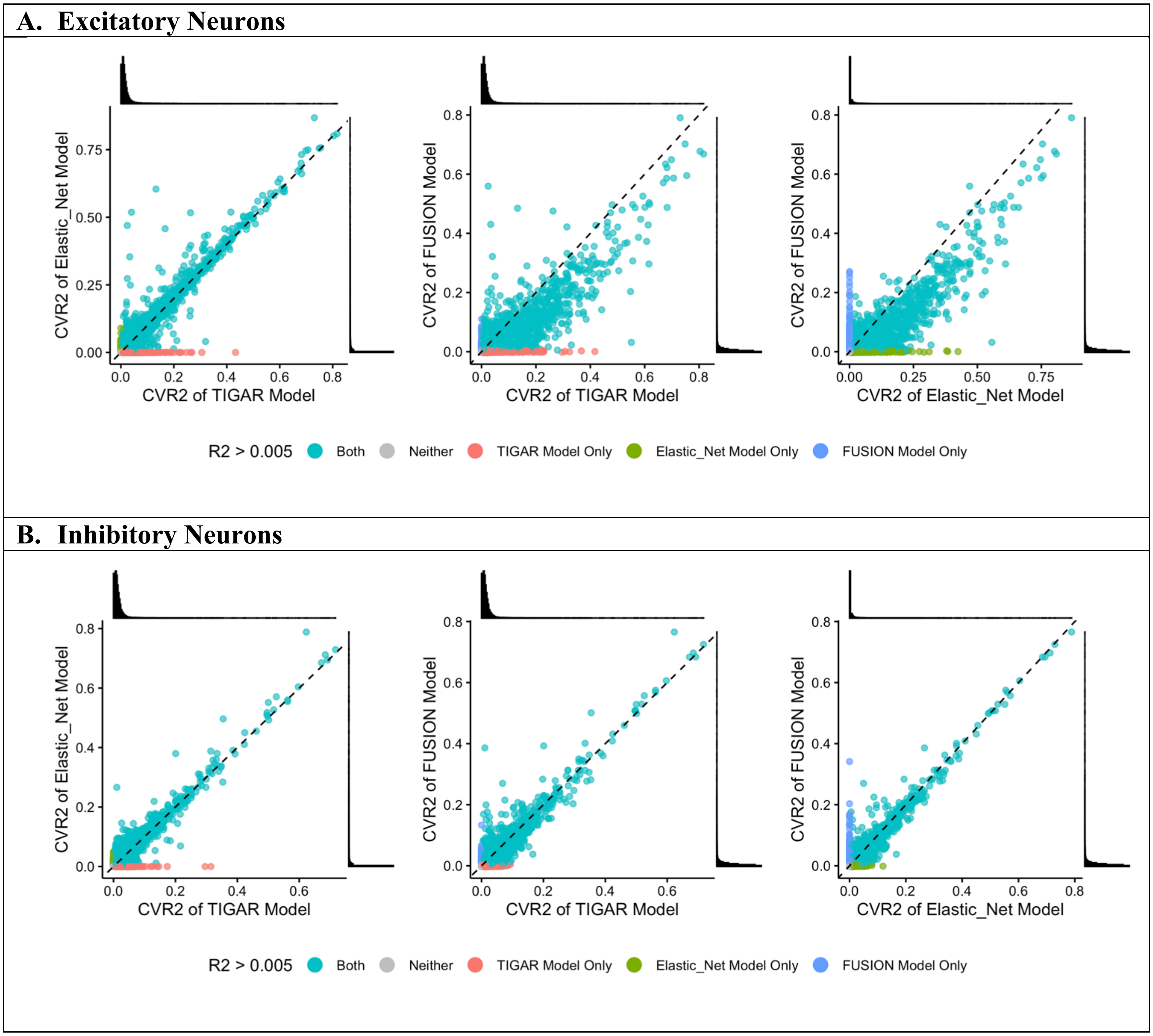

**Fig. S3. Pairwise comparison of CV $R^2$ of cell-type-specific gene expression imputation models by TIGAR/DPR, PrediXcan/EN, and FUSION/BestModel in Oligodendrocytes and Oligodendrocyte Precursor Cells.**
Each panel presents scatter plots comparing CV $R^2$ between pair-wise models, with each dot representing a gene. Colors indicate different gene groups: genes with CV $R^2$ > 0.5% in both compared models (Both:teal), in only one of the two compared models (TIGAR-only:red, Elastic Net-only:green, or FUSION-only:blue), or in neither models (Neither:gray).

**A. Oligodendrocytes**

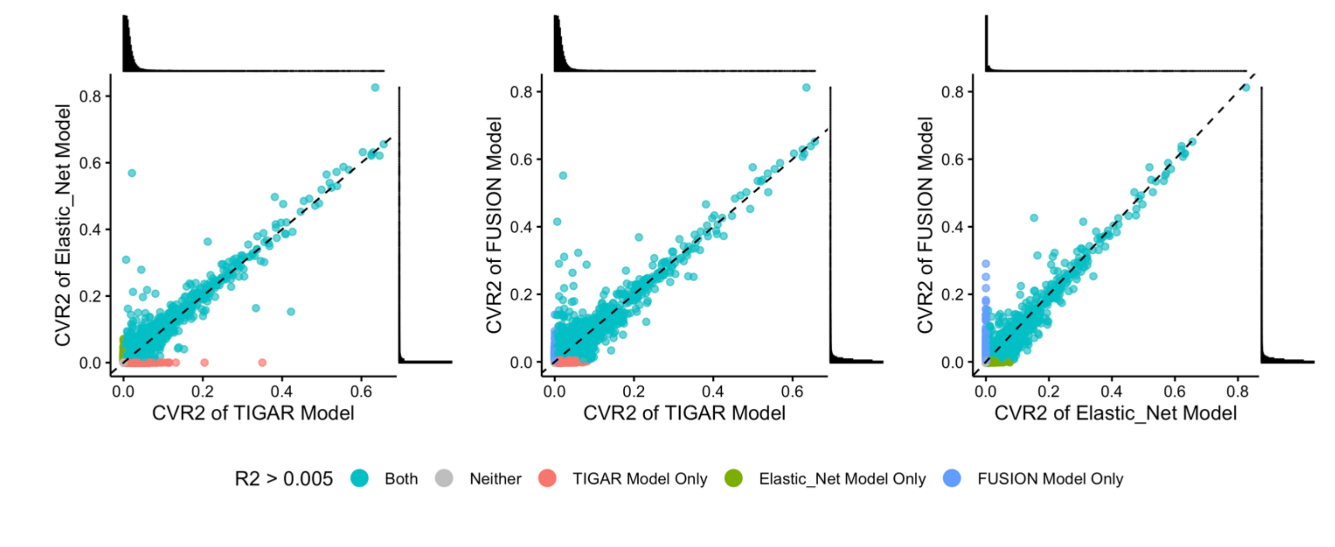


**B. Oligodendrocyte Precursor Cells**

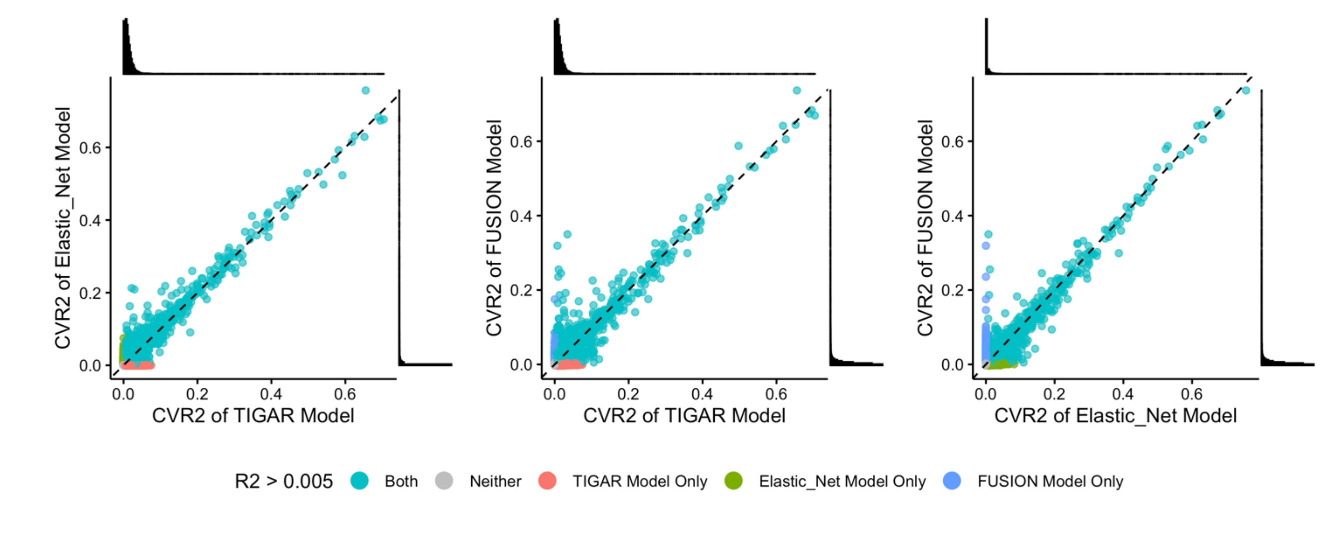

**Fig. S4. Pairwise comparison of CV $R^2$ of protein abundance imputation models by TIGAR/DPR, PrediXcan/EN, and FUSION/BestModel.**

Each panel presents scatter plots comparing CV $R^2$ between pair-wise models, with each dot representing a gene. Colors indicate different gene groups: genes with CV $R^2 > 0.5\%$ in both compared models (Both:teal), in only one of the two compared models (TIGAR-only:red, Elastic Net-only:green, or FUSION-only:blue), or in neither models (Neither:gray).

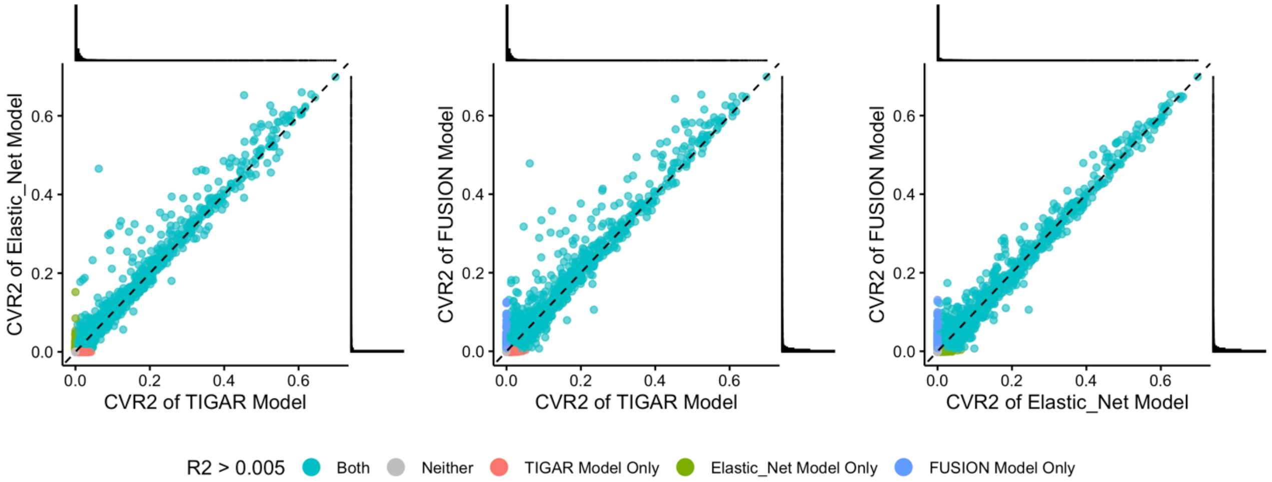

**Fig. S5. Quantile-Quantile (Q-Q) plots of transcriptome-wide -log10(p-values) obtained by TWAS-O and three individual tools for studying AD dementia in astrocytes.**
Inflated false positive rates are observed in all TWAS results.

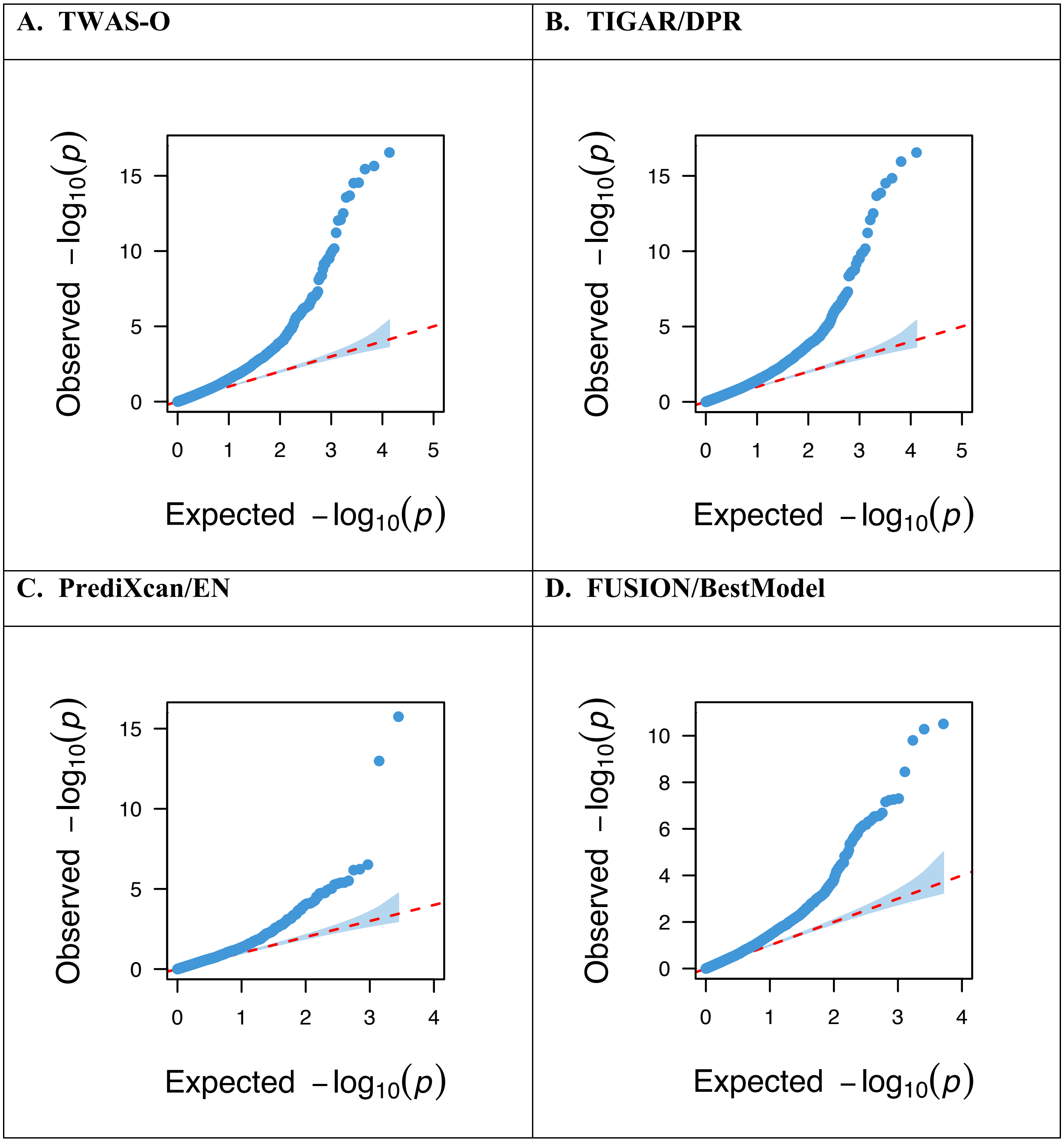

**Fig. S6. Quantile-Quantile (Q-Q) plots of transcriptome-wide -log10(p-values) obtained by TWAS-O and three individual tools for studying AD dementia in microglia.**
Inflated false positive rates are observed in all TWAS results.

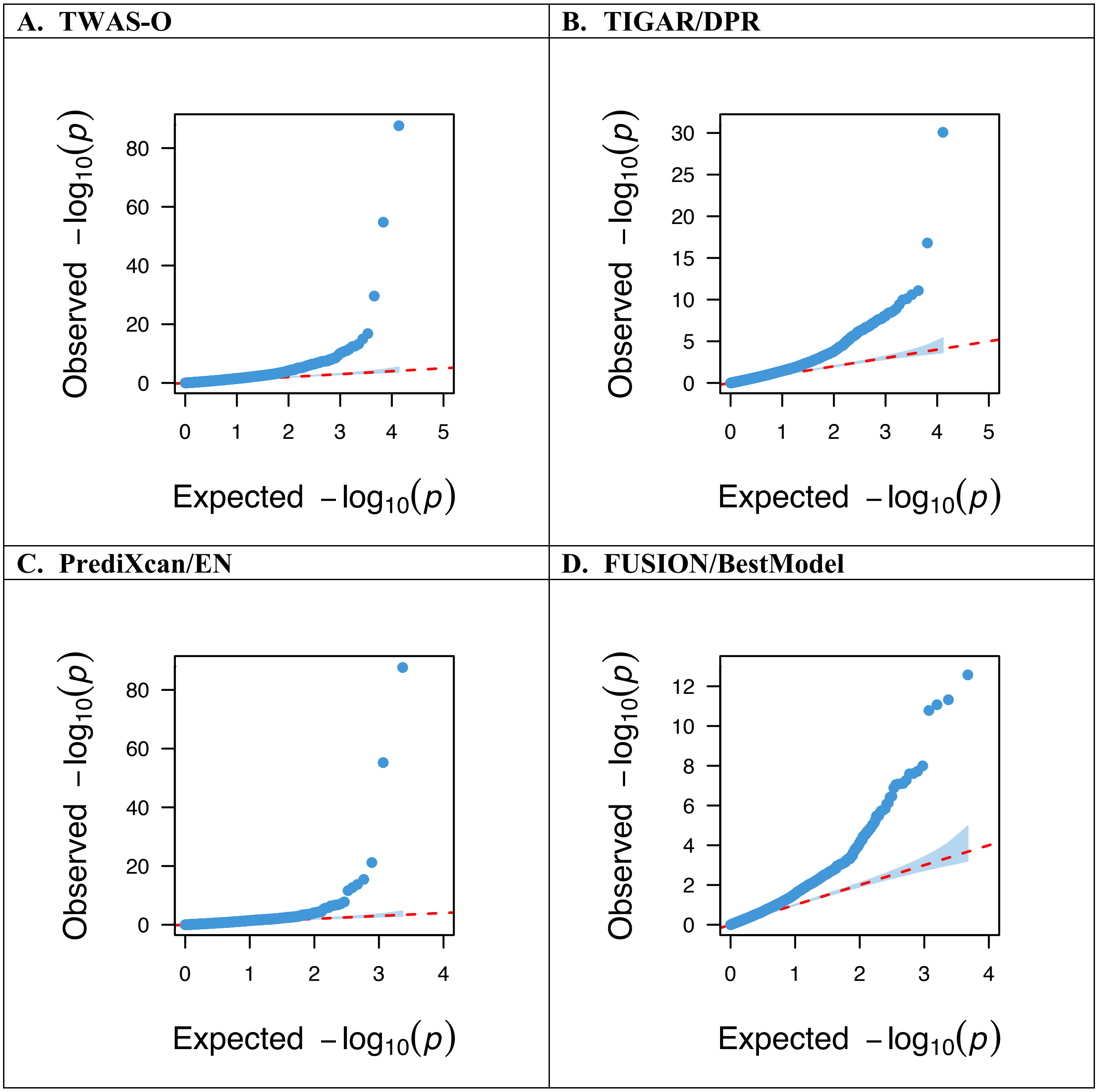

**Fig. S7. Quantile-Quantile (Q-Q) plots of transcriptome-wide -log10(p-values) obtained by TWAS-O and three individual tools for studying AD dementia in excitatory neurons.** Inflated false positive rates are observed in all TWAS results.

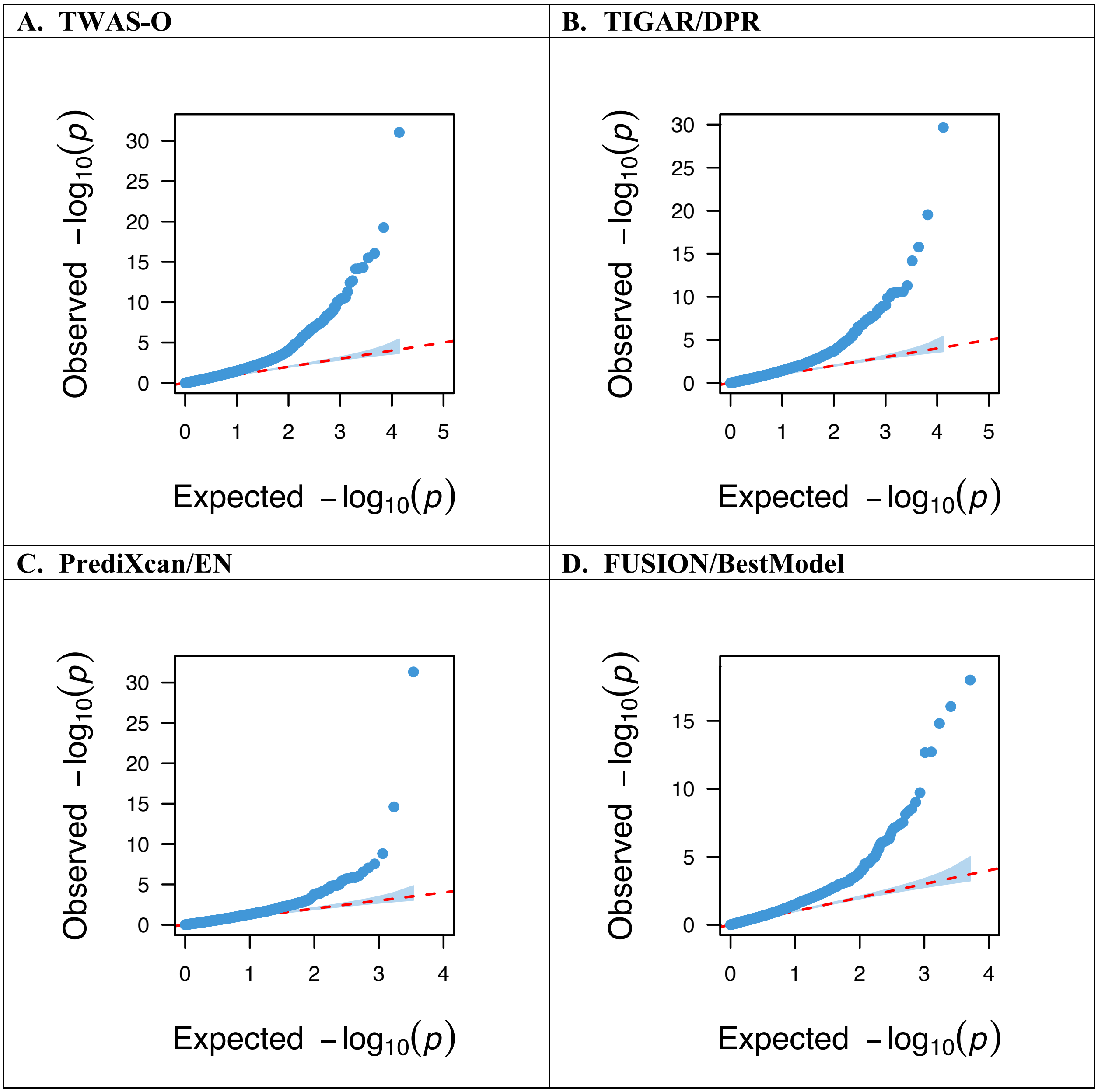

**Fig. S8. Quantile-Quantile (Q-Q) plots of transcriptome-wide -log10(p-values) obtained by TWAS-O and three individual tools for studying AD dementia in inhibitory neurons.** Inflated false positive rates are observed in all TWAS results.

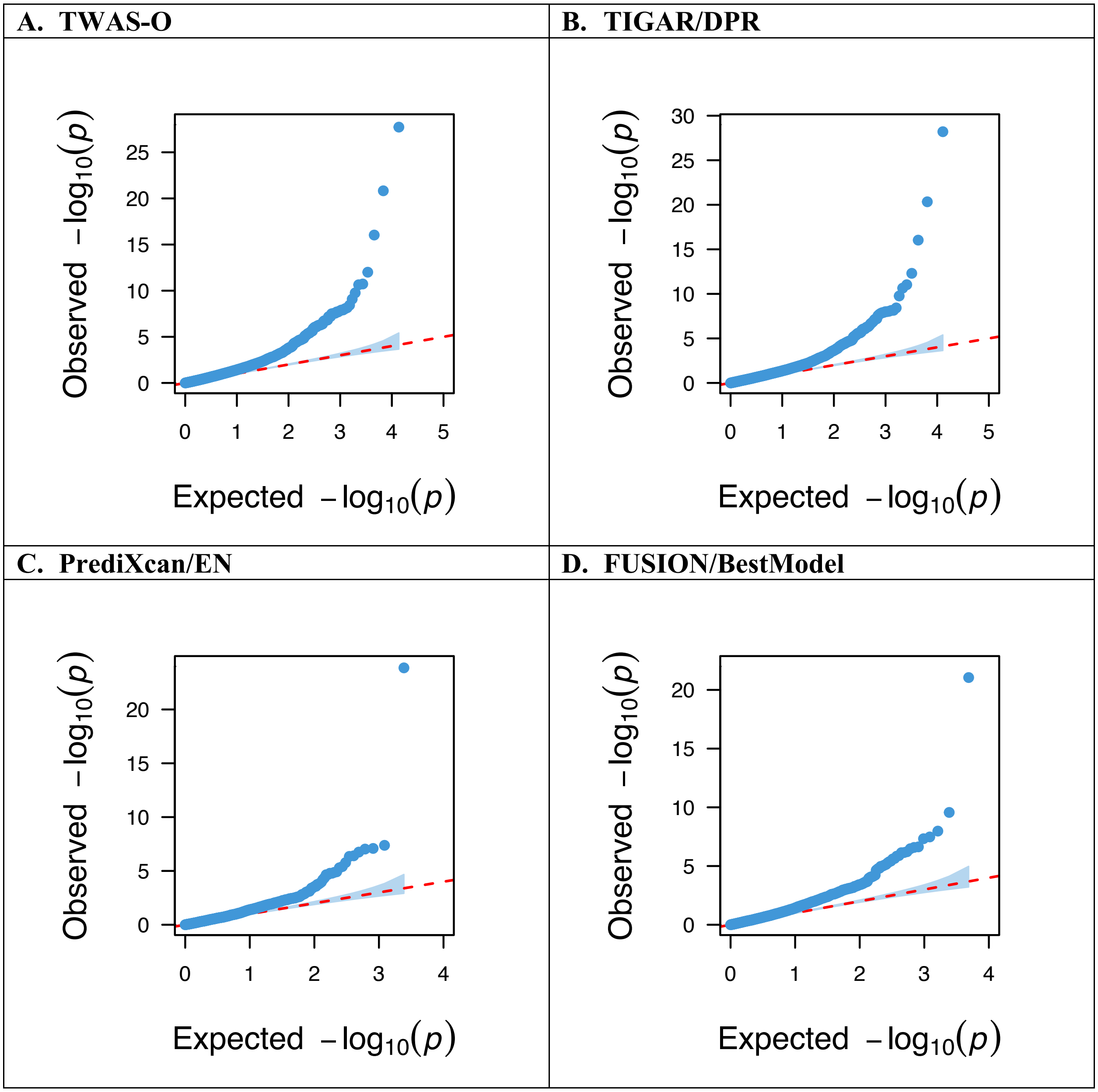

**Fig. S9. Quantile-Quantile (Q-Q) plots of transcriptome-wide -log10(p-values) obtained by TWAS-O and three individual tools for studying AD dementia in oligodendrocytes.** Inflated false positive rates are observed in all TWAS results.

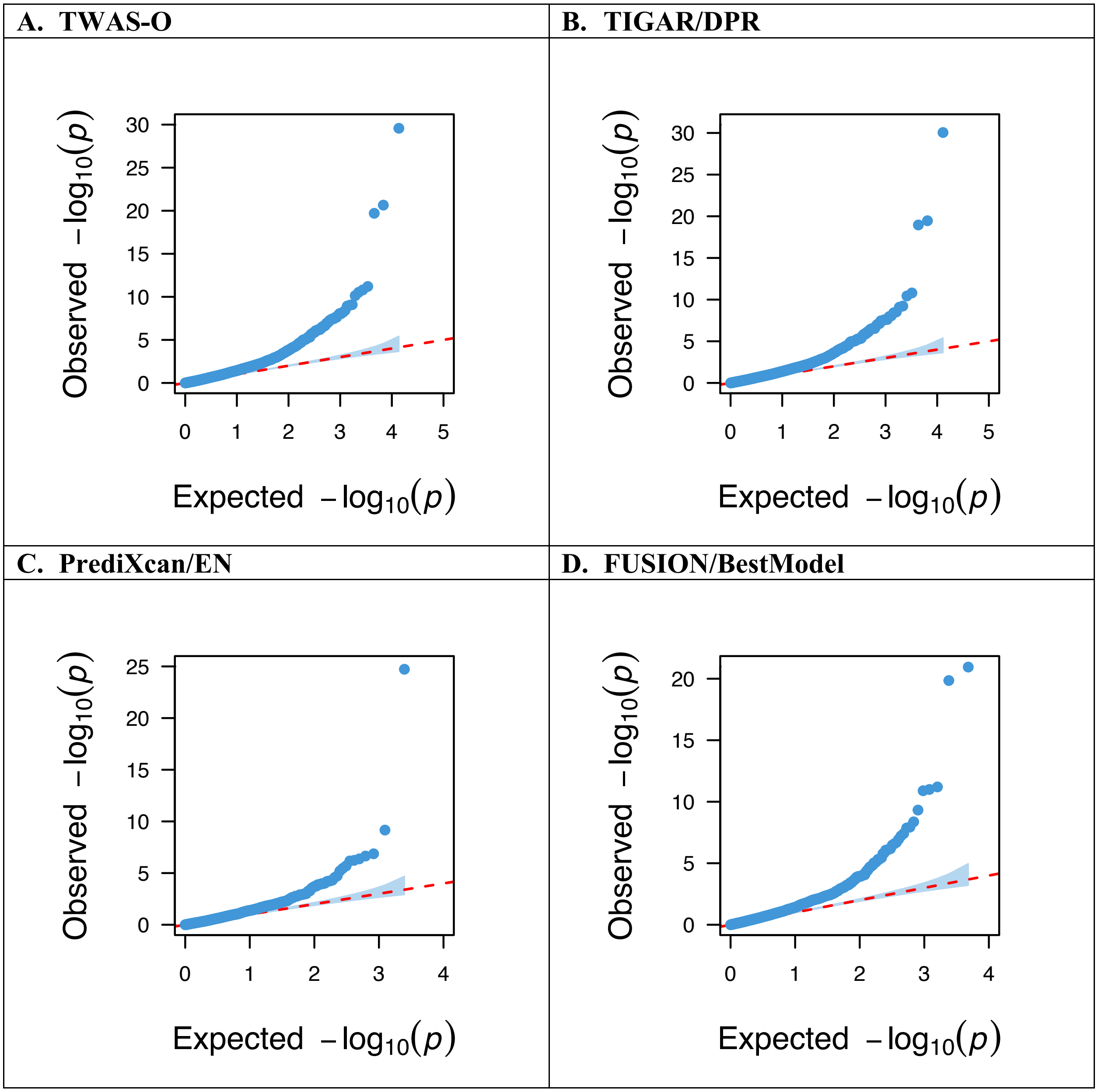

**Fig. S10. Quantile-Quantile (Q-Q) plots of transcriptome-wide -log10(p-values) obtained by TWAS-O and three individual tools for studying AD dementia in oligodendrocyte precursor cells.**

Inflated false positive rates are observed in all TWAS results.

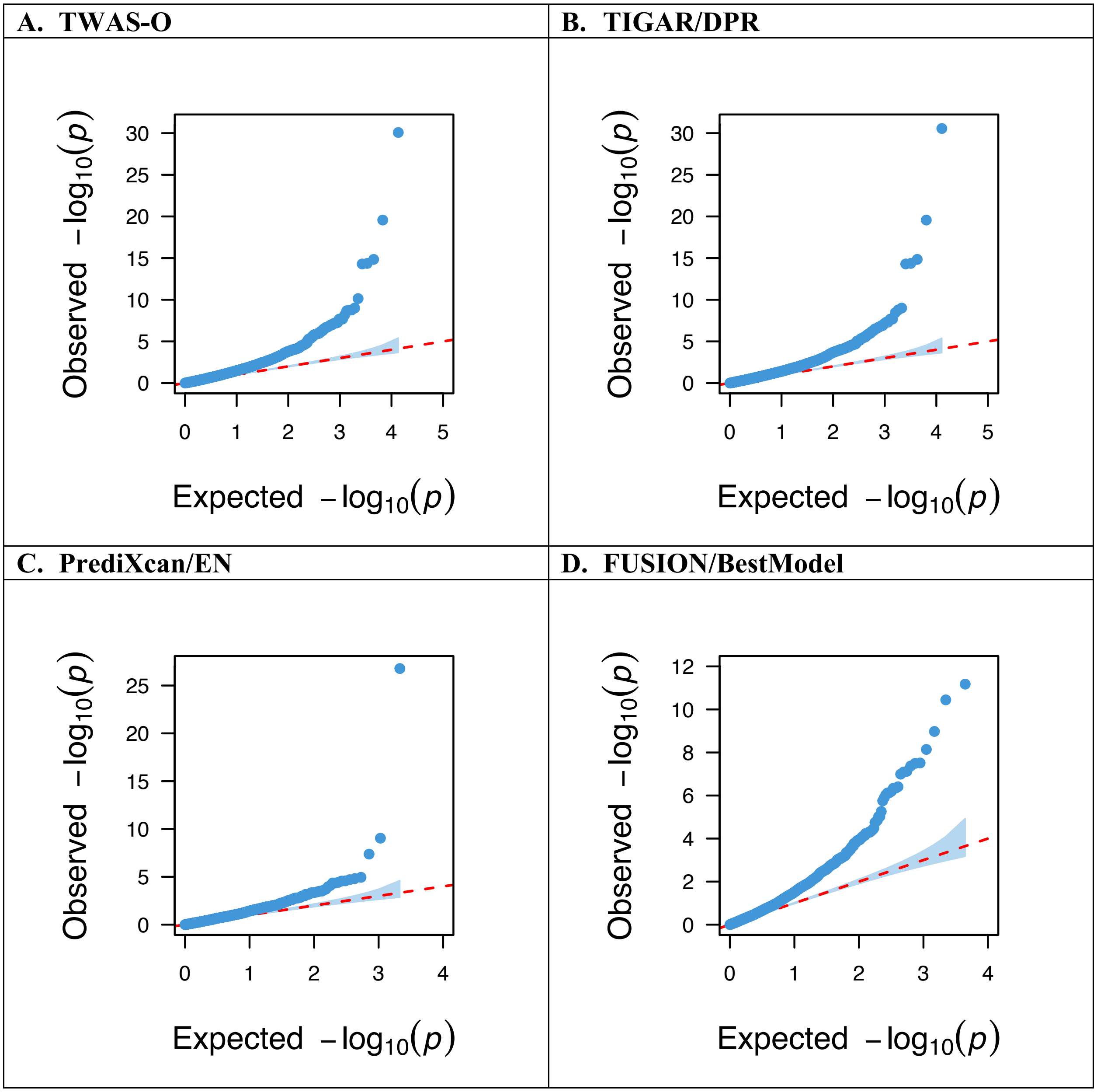

**Fig. S11. Quantile-Quantile (Q-Q) plots of proteome-wide -log10(p-values) obtained by PWAS-O and three individual tools for studying AD dementia.**
Inflated false positive rates are observed in all PWAS results.

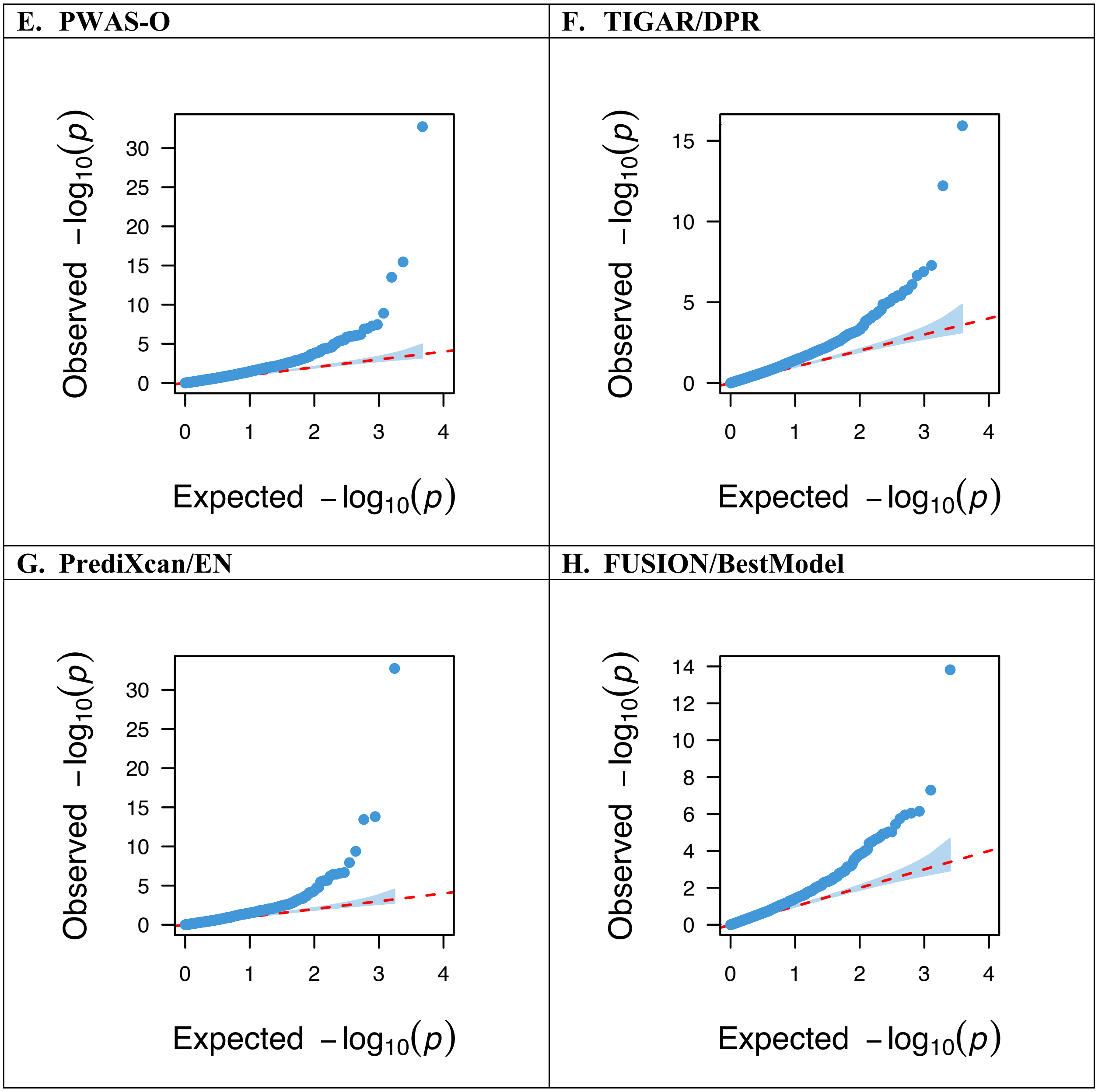

**Fig. S12. Locus plots of the fine-mapped results by GIFT of TWAS-O results in astrocytes (Ast).**

Each panel represents the fine-mapped results by GIFT within one genomic region (±1Mb around the top significant TWAS-O risk genes) that contains multiple significant TWAS-O risk genes. X-axis: chromosomal position (Mb); Y-axis: -log10(p-values) by GWAS (gray dots), TWAS-O (blue squares), and GIFT (red diamonds); Dashed horizontal line: the significance threshold of 0.05 for GIFT p-value. Genes with significant GIFT p-values < 0.05 are labeled with their gene names.

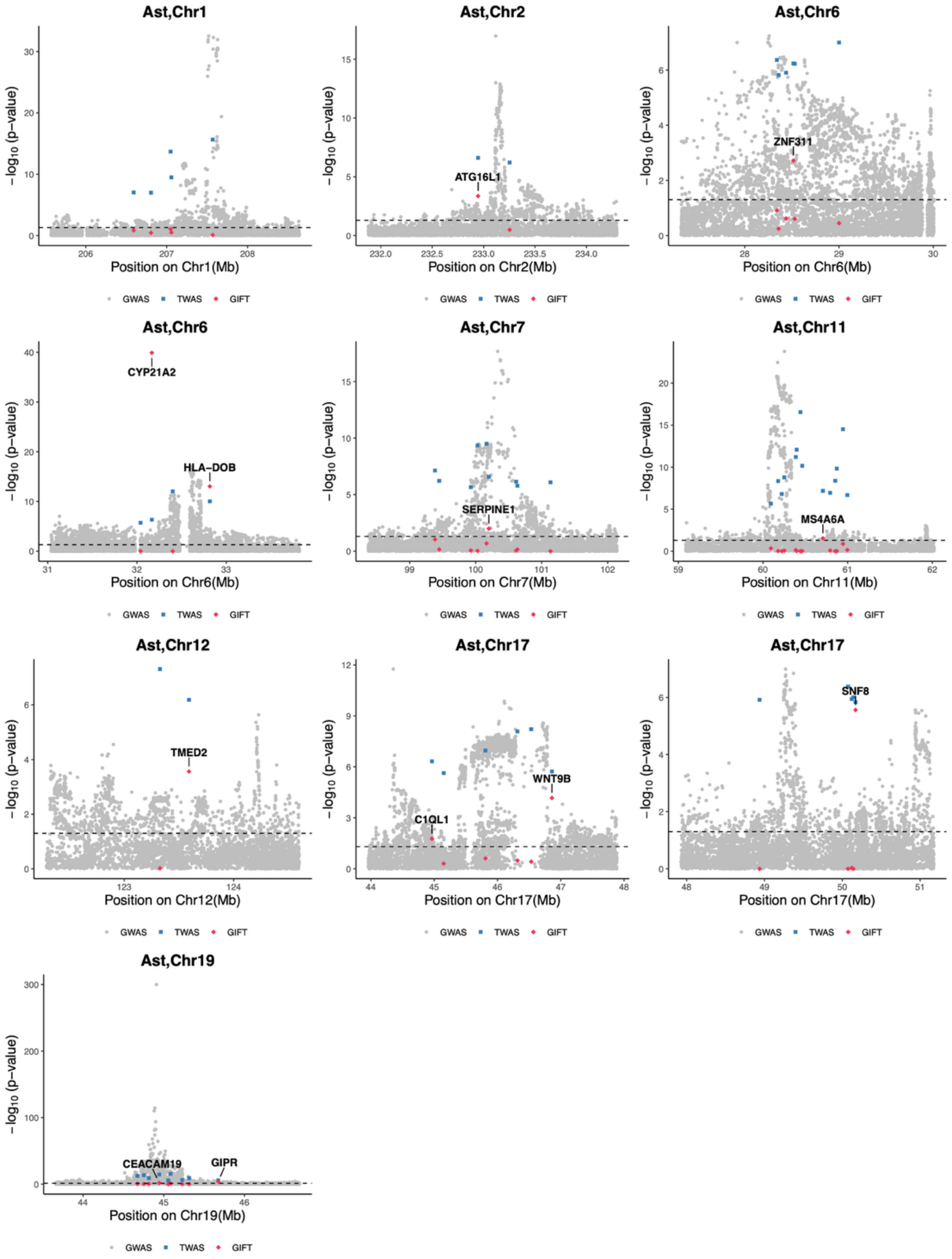

**Fig. S13. Locus plots of the fine-mapped results by GIFT of TWAS-O results in microglia (Mic).**

Each panel represents the fine-mapped results by GIFT within one genomic region (±1Mb around the top significant TWAS-O risk genes) that contains multiple significant TWAS-O risk genes. X-axis: chromosomal position (Mb); Y-axis: -log10(p-values) by GWAS (gray dots), TWAS-O (blue squares), and GIFT (red diamonds); Dashed horizontal line: the significance threshold of 0.05 for GIFT p-value. Genes with significant GIFT p-values < 0.05 are labeled with their gene names.

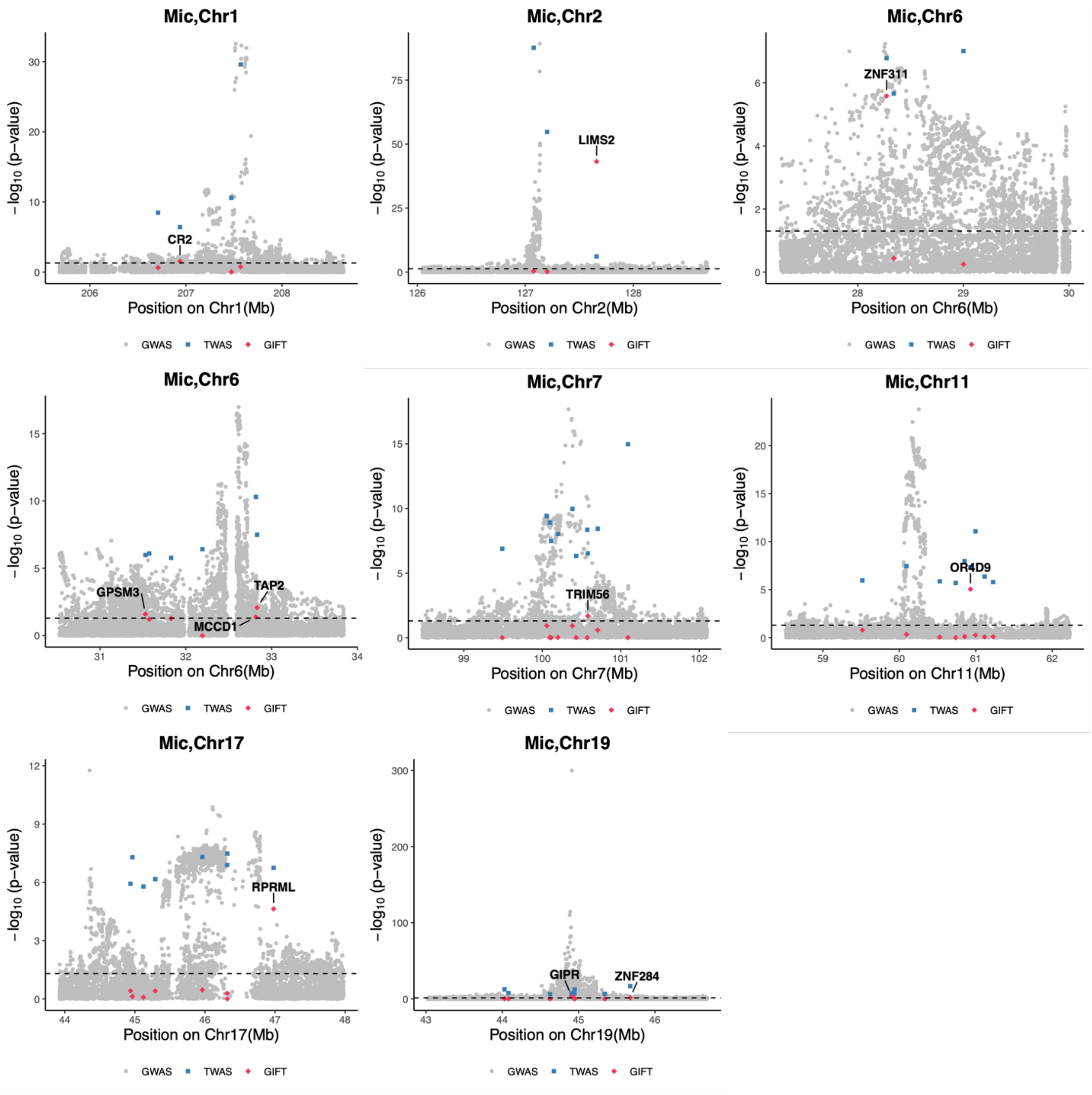

**Fig. S14. Locus plots of the fine-mapped results by GIFT of TWAS-O results in excitatory neurons (Ex).**

Each panel represents the fine-mapped results by GIFT within one genomic region (±1Mb around the top significant TWAS-O risk genes) that contains multiple significant TWAS-O risk genes. X-axis: chromosomal position (Mb); Y-axis: -log10(p-values) by GWAS (gray dots), TWAS-O (blue squares), and GIFT (red diamonds); Dashed horizontal line: the significance threshold of 0.05 for GIFT p-value. Genes with significant GIFT p-values < 0.05 are labeled with their gene names.

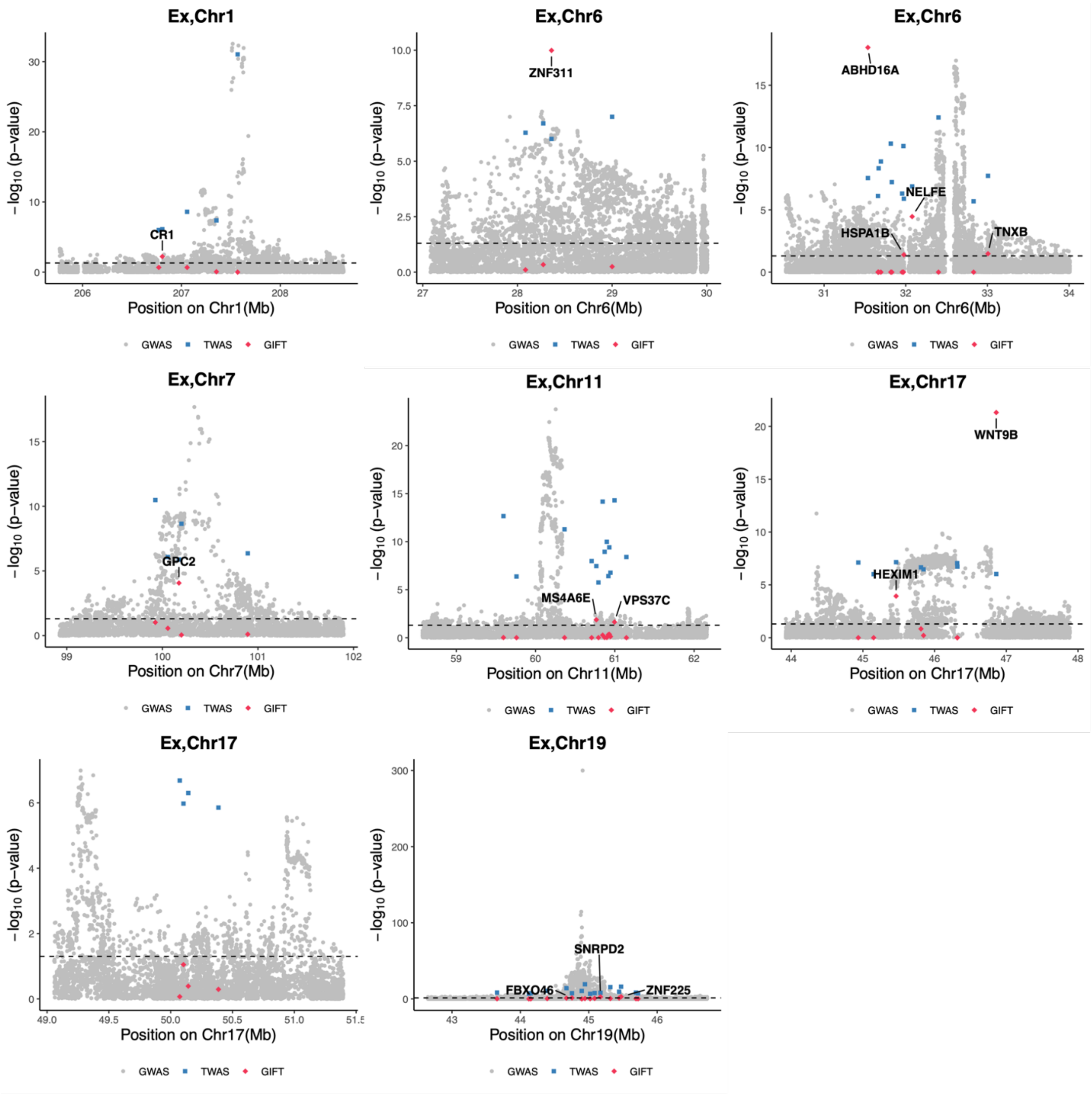

**Fig. S15. Locus plots of the fine-mapped results by GIFT of TWAS-O results in inhibitory neurons (In).**

Each panel represents the fine-mapped results by GIFT within one genomic region (±1Mb around the top significant TWAS-O risk genes) that contains multiple significant TWAS-O risk genes. X-axis: chromosomal position (Mb); Y-axis: -log10(p-values) by GWAS (gray dots), TWAS-O (blue squares), and GIFT (red diamonds); Dashed horizontal line: the significance threshold of 0.05 for GIFT p-value. Genes with significant GIFT p-values < 0.05 are labeled with their gene names.

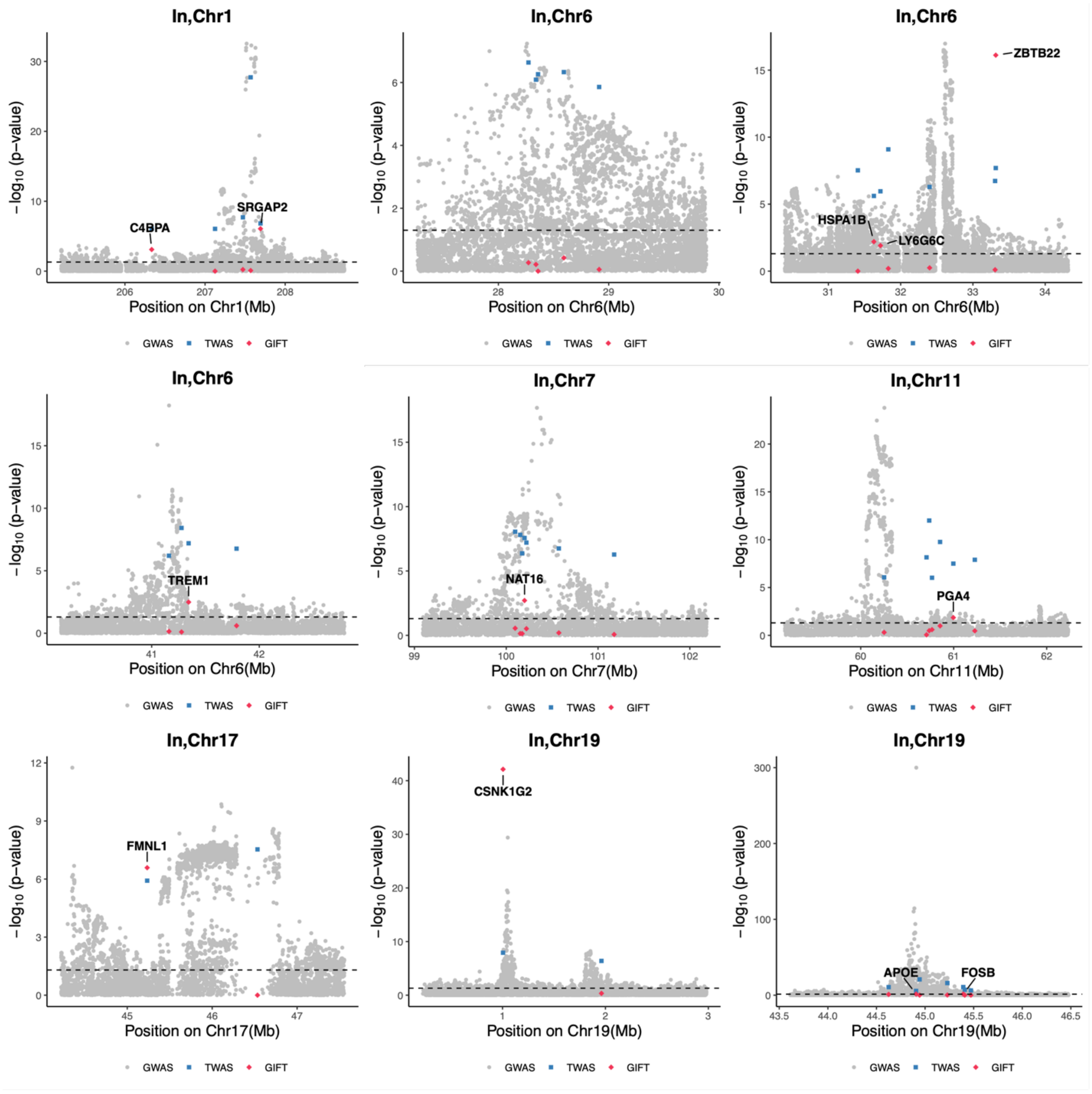

**Fig. S16. Locus plots of the fine-mapped results by GIFT of TWAS-O results in oligodendrocytes (Oli).**

Each panel represents the fine-mapped results by GIFT within one genomic region (±1Mb around the top significant TWAS-O risk genes) that contains multiple significant TWAS-O risk genes. X-axis: chromosomal position (Mb); Y-axis: -log10(p-values) by GWAS (gray dots), TWAS-O (blue squares), and GIFT (red diamonds); Dashed horizontal line: the significance threshold of 0.05 for GIFT p-value. Genes with significant GIFT p-values < 0.05 are labeled with their gene names.

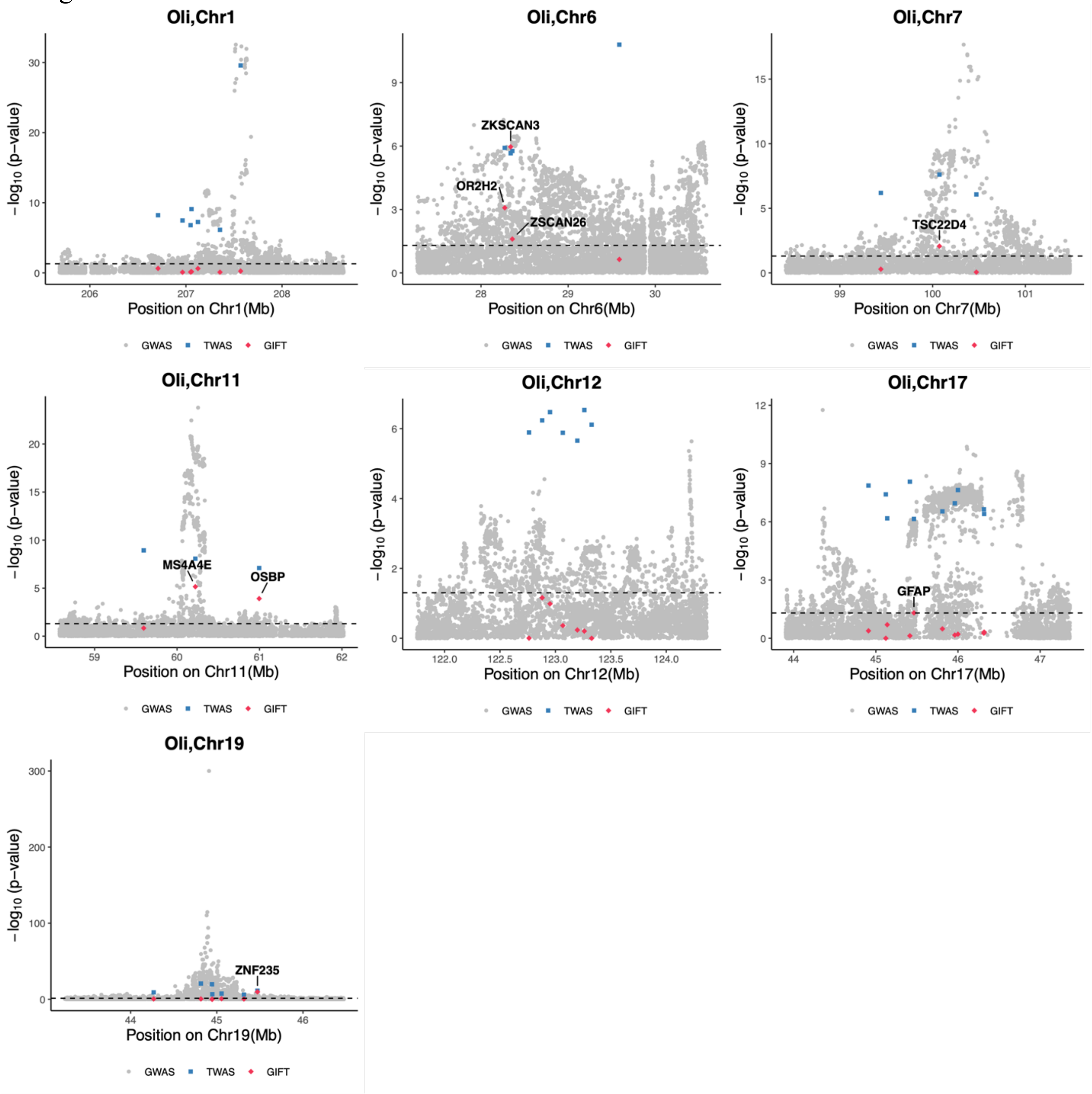

**Fig. S17. Locus plots of the fine-mapped results by GIFT of TWAS-O results in oligodendrocyte precursor cells (Opc).**
Each panel represents the fine-mapped results by GIFT within one genomic region (±1Mb around the top significant TWAS-O risk genes) that contains multiple significant TWAS-O risk genes. X-axis: chromosomal position (Mb); Y-axis: -log10(p-values) by GWAS (gray dots), TWAS-O (blue squares), and GIFT (red diamonds); Dashed horizontal line: the significance threshold of 0.05 for GIFT p-value. Genes with significant GIFT p-values < 0.05 are labeled with their gene names.

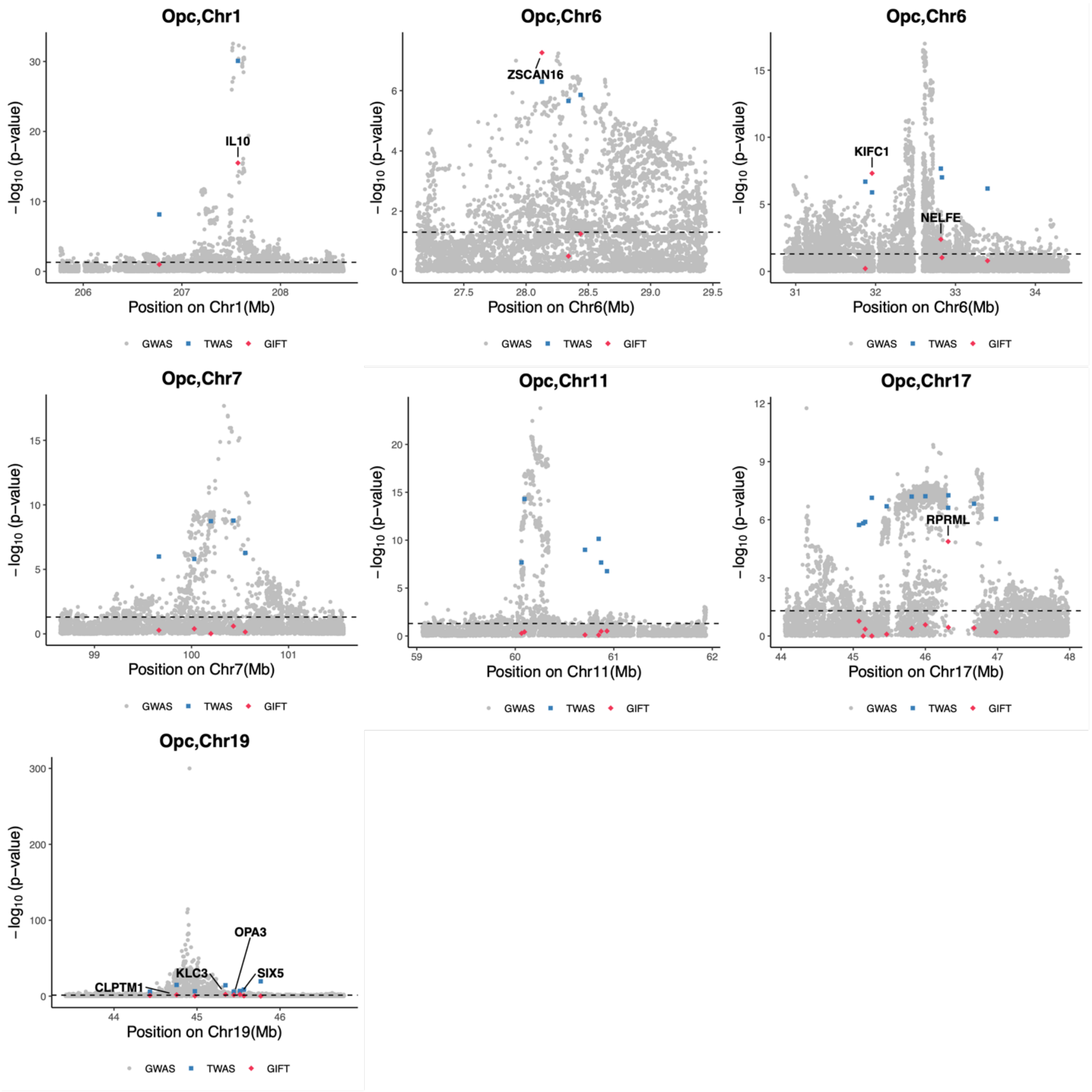

**Fig. S18. Locus plots of the fine-mapped results by GIFT of TWAS-O results in proteomics (Proteome).**

Each panel represents the fine-mapped results by GIFT within one genomic region (±1Mb around the top significant TWAS-O risk genes) that contains multiple significant TWAS-O risk genes. X-axis: chromosomal position (Mb); Y-axis: -log10(p-values) by GWAS (gray dots), TWAS-O (blue squares), and GIFT (red diamonds); Dashed horizontal line: the significance threshold of 0.05 for GIFT p-value. Genes with significant GIFT p-values < 0.05 are labeled with their gene names.

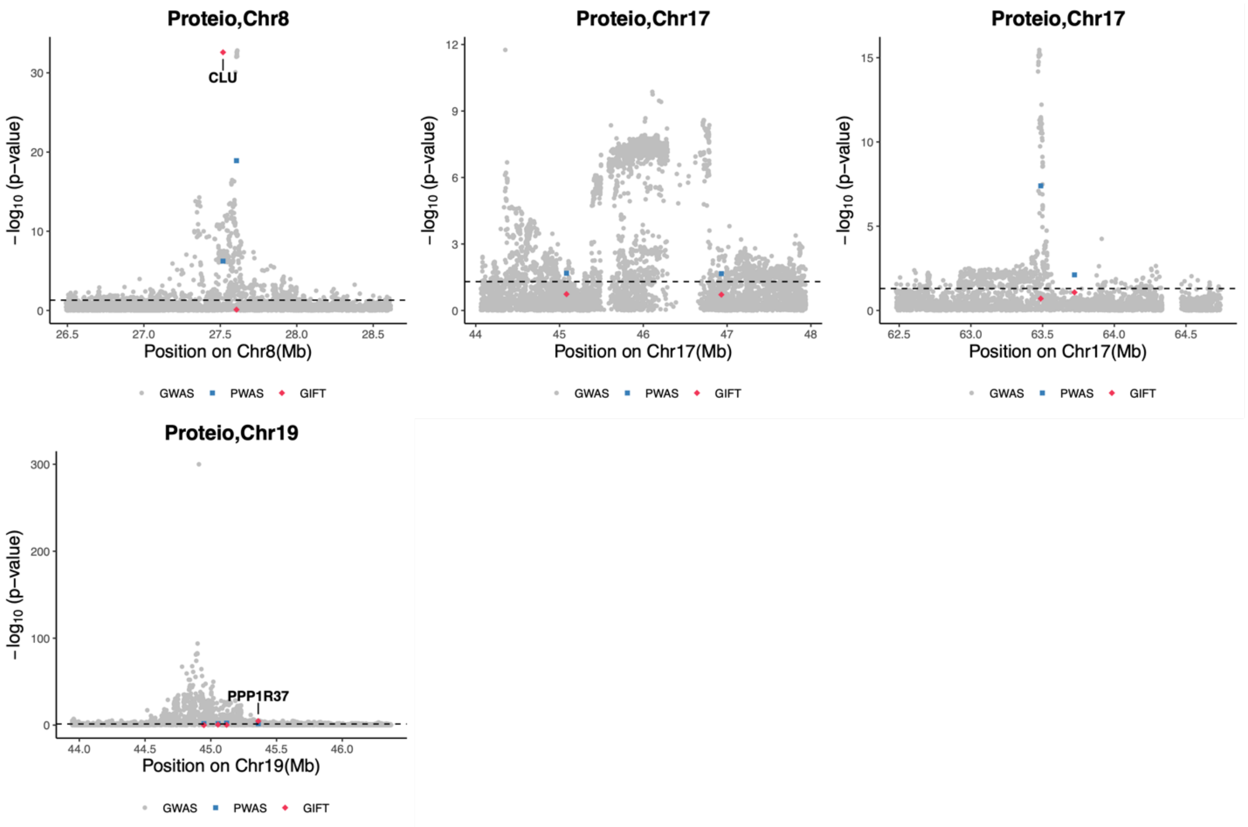

**Fig. S19. Jaccard similarity indexes of overlapped transcriptome-wide and proteome-wide significant TWAS-O/PWAS-O risk genes by pair-wise comparisons.**
Jaccard similarity index is the proportion of overlapped significant risk genes among the union of unique significant risk genes detected by both compared analyses. Only genes with exactly the same names are considered overlapped in the calculation of Jaccard similarity indexes. Higher Jaccard Index value represents higher proportions of overlapped findings.

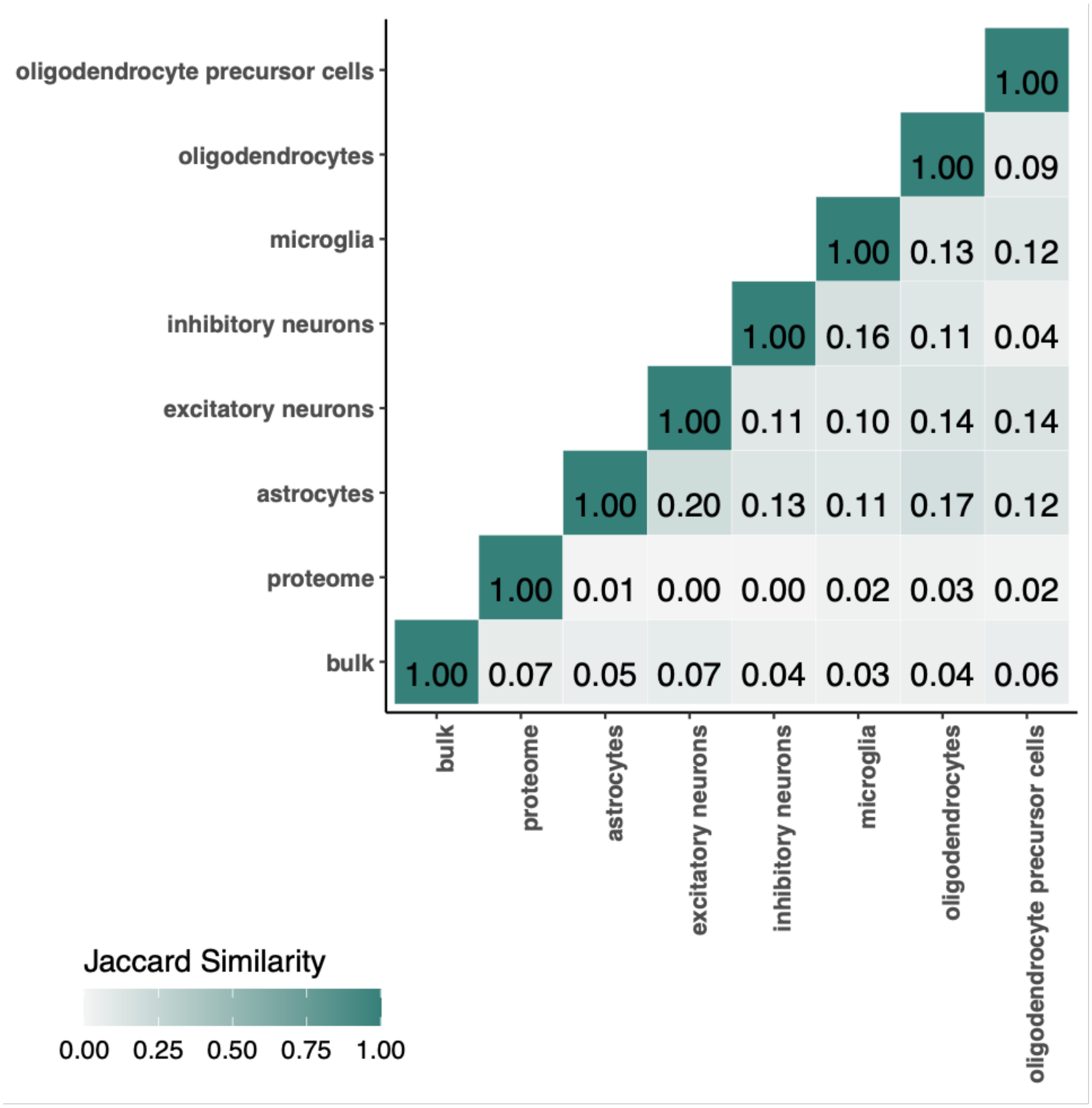
